# Genome-wide analyses of early-onset acute myocardial infarction identify 29 novel loci by whole genome sequencing

**DOI:** 10.1101/2022.05.22.22275428

**Authors:** Yeonsu Jeon, Sungwon Jeon, Whan-Hyuk Choi, Kyungwhan An, HanSol Choi, Byoung-Chul Kim, Weon Kim, Sang Yeob Lee, Jang-Whan Bae, Jin-Yong Hwang, Min Gyu Kang, Yeonkyung Kim, Younghui Kang, Yeo Jin Kim, Byung Chul Kim, Jong Bhak, Eun-Seok Shin

## Abstract

Early-onset acute myocardial infarction (AMI) may have a higher genetic predisposition than late-onset AMI does. The present study aimed to identify and characterize germline variants that affect early-onset AMI using whole-genome sequencing (WGS). We performed a genome-wide association study based on WGS of 1,239 Koreans, including 596 early-onset AMI patients and 643 healthy individuals. Patients with AMI who underwent percutaneous coronary intervention (PCI) caused by atherothrombotic occlusive lesions were included in the study. A total of 29 novel loci were found to be associated with early-onset AMI. These loci are involved in thrombosis, fibrinolysis, inflammation, and lipid metabolism. One of the associated single nucleotide variants (SNVs), rs1614576, located upstream of *PRKCB*, is known to be associated with thrombus formation. Additionally, the results revealed a novel locus, rs78631167, located upstream of *PLAUR* which plays a critical role in regulating plasminogen activation and is related to fibrinolysis. The association between early-onset AMI and rs9357455, which is located upstream of *PHACTR1* that regulates inflammation in AMI, was found. Moreover, we could identify a lipid metabolism related genetic risk locus, rs5072, in the *APOA1-AS* gene. This study provides new evidence supporting the genetic association between early-onset AMI and thrombosis and fibrinolysis, as well as inflammation and lipid metabolism, by analyzing the whole-genome of 596 patients with early-onset AMI who have been treated with PCI. Our findings highlight potential genetic markers for the prediction and management of AMI, as well as for understanding the etiology of AMI.

## INTRODUCTION

Acute myocardial infarction (AMI) is a leading cause of death with more than one million deaths per year worldwide (1). The majority of AMI cases occur in individuals >65 years of age. Early-onset AMI is rare with a prevalence of 1-5%, and genetic markers discovered from them are probably more strongly associated than the late-onset counterpart (2-4). Until now, many AMI studies have found loci associated with lipid and cholesterol metabolism which is associated with atherosclerotic plaque development and progression (2, 5-8). Atherosclerotic plaque causes coronary artery disease; however, a thrombus consisting of platelet aggregates and fibrin strands ultimately leads to the clinical manifestations of AMI (2, 5, 9, 10). However, the genetic factors that contribute to atherothrombotic coronary occlusion leading to AMI are not clearly understood (11). In addition, recent clinical studies have shown that the occurrence of AMI may be caused by an imbalance between thrombosis and fibrinolysis, a conflicting mechanism (12). Fibrinolysis is involved in plaque healing and has emerged as a critical mechanism of AMI (12). However, few studies have been conducted on the genetic factors contributing to AMI by fibrinolysis (13).

The present study aimed to identify novel genomic loci associated with atherothrombotic coronary occlusion, leading to the clinical manifestations of AMI. We report a genome-wide association study (GWAS) based on whole-genome sequencing (WGS) in patients with early-onset AMI who were eventually treated with percutaneous coronary intervention (PCI), and patients with non-obstructive AMI were excluded.

## RESULTS

### Novel Genomic Loci Associated with Early-Onset AMI

Twenty-nine novel genomic loci (85 suggestive variants) for early-onset AMI were identified using WGS-based GWAS over the genome-wide suggestive threshold (*P*-value cutoff = 1 × 10^−5^; Fig. 1, Table 1, 2, and Supplementary Material, Fig. S1, S2, Table S1). Approximately 86% of the novel loci were not covered in SNP chips such as Illumina human exome beadchip, Humanhap 610 Quad Chip, and Affymetrix 6.0 GeneChip (Supplementary Material, Table S2 and S3) (2, 5, 14). The 29 novel loci included loci that are known to be related to thrombosis and fibrinolysis. Locus rs1614576, located upstream of protein kinase C beta (*PRKCB*) gene, had a suggestive association with early-onset AMI (OR = 1.646, *P* = 3.41 × 10^−6^; Fig. 2A). It is involved in platelet responses implicated in thrombus formation (15). Another novel locus, rs78631167 (OR = 0.473, *P* = 2.93 × 10^−6^) located upstream of the *PLAUR* (plasminogen activator, urokinase receptor), was identified (Fig. 2B), and it is involved in fibrinolysis (16). This locus was predicted to be located in a regulatory region with a RegulomeDB rank of 4 (17), representing transcription factor (TF) binding and DNase peak site. *PLAUR* encodes CD87, which binds pro-urokinase to the cell surface and converts plasminogen to plasmin, resulting in clot lysis (18). CD87 insufficiency results in impaired fibrinolysis and reduced clot dissolution (18). In addition, a novel locus related to inflammation, rs9357455, located upstream of the *PHACTR1* (phosphatase and actin regulator 1) was also identified, suggesting an association with early-onset AMI (OR = 0.666, *P* = 9.73 × 10^−6^; Fig. 2C). This locus was predicted to have regulatory functions, with a RegulomeDB rank of 5 (17), including regulation of oxidative stress and inflammation in coronary artery endothelial cells via interactions with NF-kB/p65 (19). Previous myocardial infarction studies have reported that variants, such as rs12526453 and rs9349379, in the *PHACTR1* gene are associated with myocardial infarction (MI) (Supplementary Material, Table S4) (2, 20). The results of this study revealed an association between early-onset AMI and a novel variant called rs9357455 in the *PHACTR1* gene (Fig. 2C). Moreover, rs5072, a genetic risk locus for early-onset AMI (OR = 1.545, *P* = 8.61 × 10^−6^) located in the intron of the *APOA1-AS* (apolipoprotein A1 antisense RNA) gene which is involved in lipid metabolism, was also identified (Fig. 2D). It regulates the expression of *APOA1* whose protein is a component of HDL cholesterol and a major participant in the regulation of reverse cholesterol transport from peripheral tissues to the liver (21). Protein-protein interaction (PPI) analysis revealed that dissolution of fibrin clot (HSA-75205), complement and coagulation cascades (hsa04610), and cholesterol metabolism (hsa04979) pathways were enriched from the genes on which the 29 novel loci are located (Supplementary Material, Fig. S3 and Table S5-S7).

**Table 1.** Baseline characteristics.

|  | Early-onset AMI (N=596) | Control (N=643) |
| --- | --- | --- |
| **Male, n (%)** | 486 (81.5) | 328 (51.0) |
| **Age, median (Q1 - Q3)** | 46 (42.0-48.0) | 44 (29.5-57.0) |
| **Body mass index, mean ± SD** | 25.5 ± 3.9 | 24.0 ± 3.4 |
| **Hypertension, n (%)** | 188 (31.5) | 90 (14.0) |
| **Diabetes mellitus, n (%)** | 118 (19.8) | 33 (5.1) |
| **Current smoking, n (%)** | 360 (60.4) | 82 (12.8) |
| **Lipid levels, mg/dL** |  |  |
| **Total cholesterol, mean ± SD** | 202.7 ± 49.2 | 179.6 ± 34.3 |
| **LDL cholesterol, mean ± SD** | 129.1 ± 43.4 | 115.8 ± 33.0 |
| **HDL cholesterol, mean ± SD** | 43.9 ± 13.0 | 57.4 ± 13.9 |
| **Triglycerides, mean ± SD** | 209.1 ± 185.1 | 115.8 ± 77.0 |

AMI, acute myocardial infarction; family history of CAD, 1st degree family history of coronary artery disease; LDL, low-density lipoprotein; HDL, high-density lipoprotein. Values are mean ± SD, median (interquartile range, 25th - 75th), or n (%).

**Table 2.**
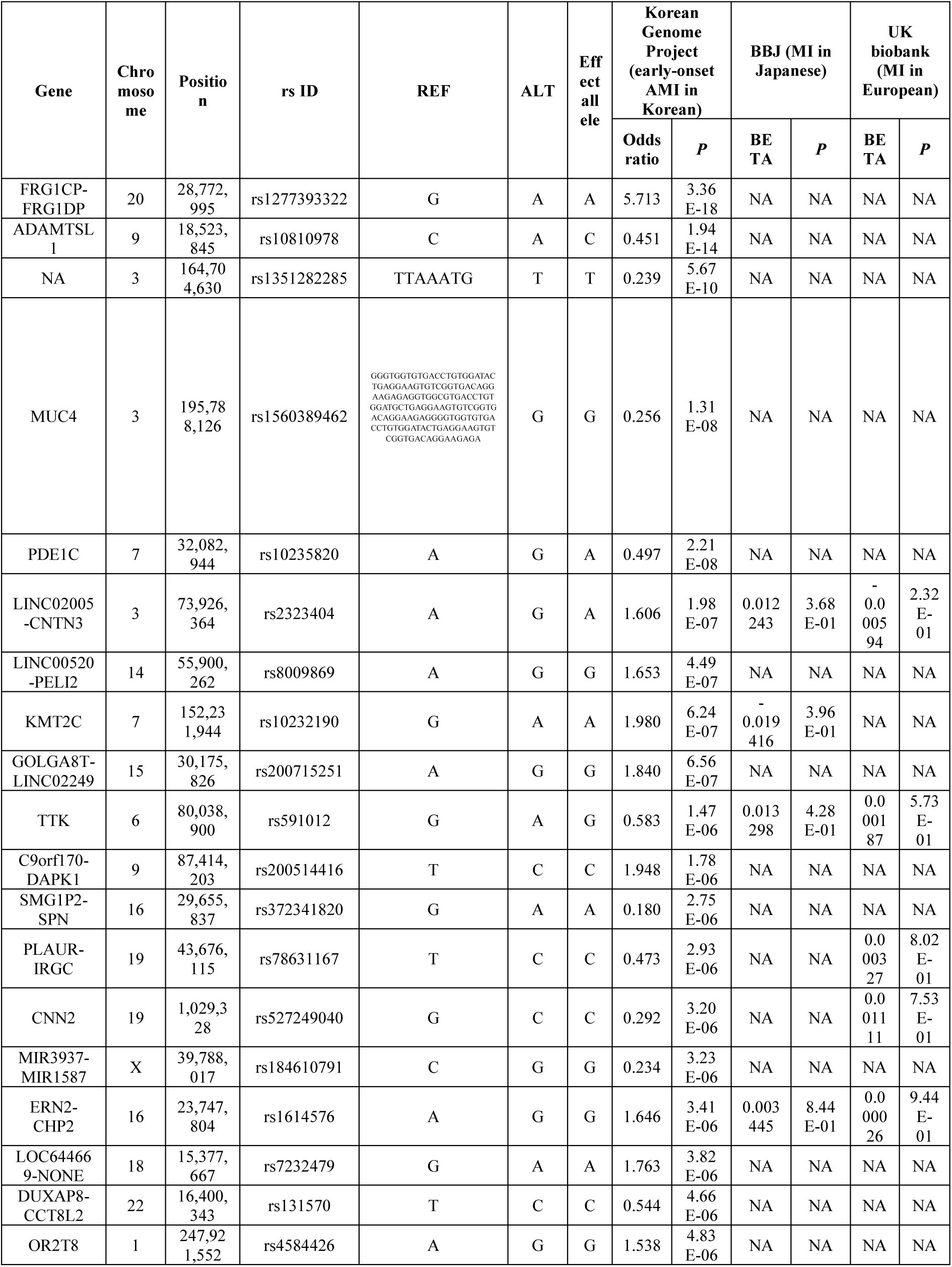
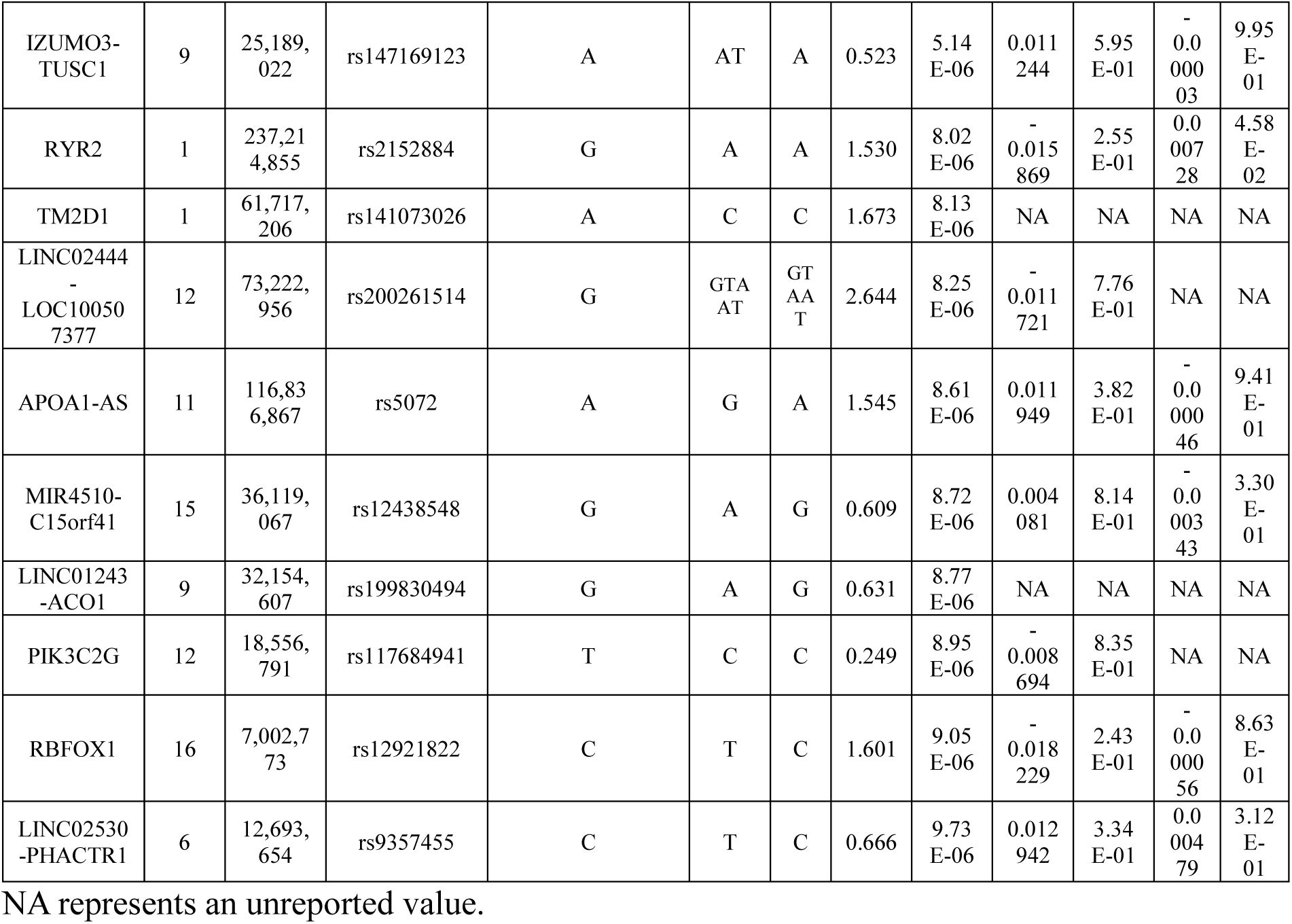
Novel genomic loci for early-onset AMI compared to European and Japanese MI studies.

**Figure 1.**
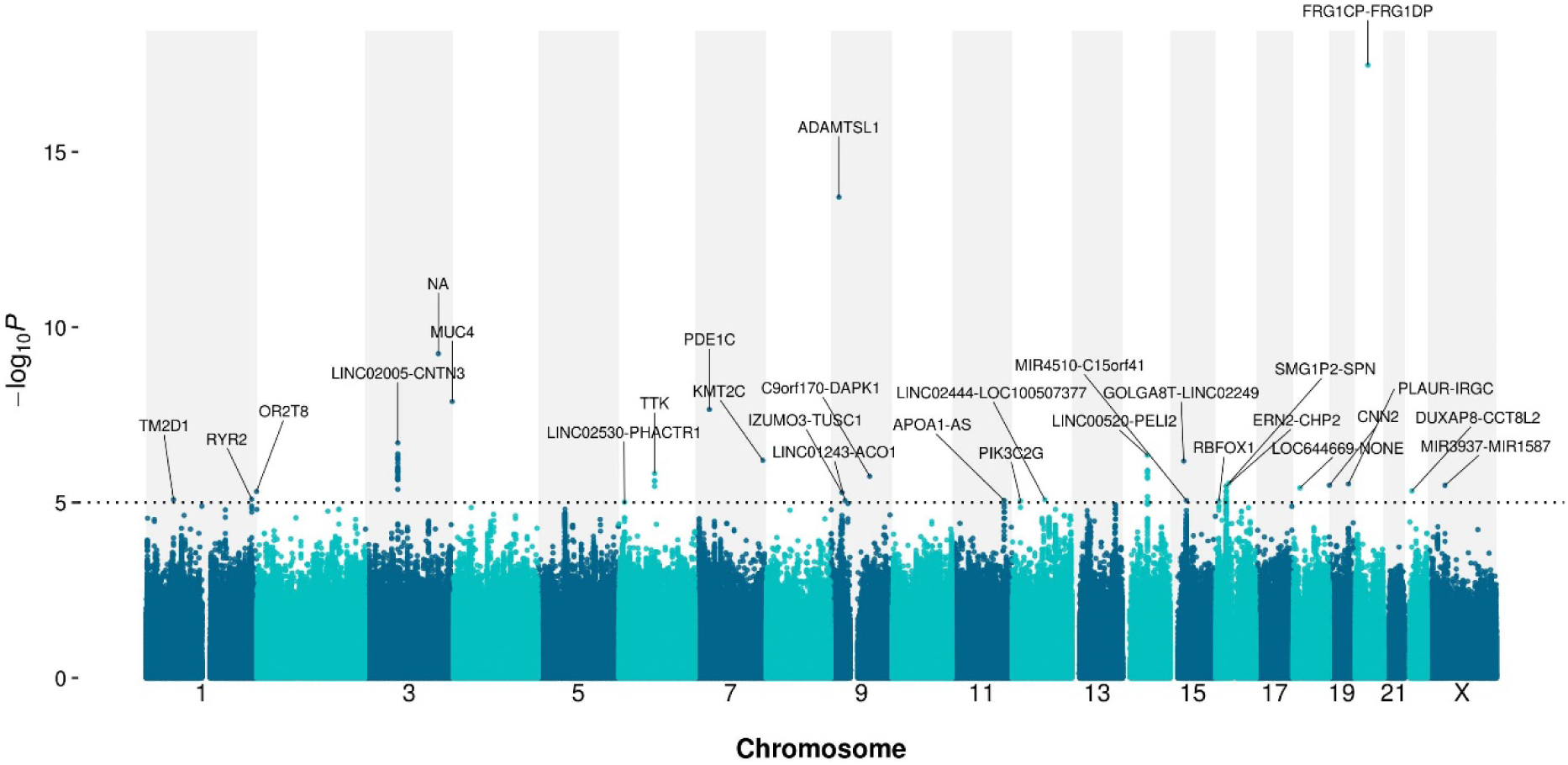
Manhattan plot of the associated genomic loci for early-onset AMI via a GWAS. X-axis indicates the genomic positions (hg38). Y-axis indicates -log_10_ *P*-value from the GWAS. The dotted line indicates the genome-wide suggestive threshold (1 × 10^−5^). The suggestive genomic loci are presented as gene symbols.

**Figure 2.**
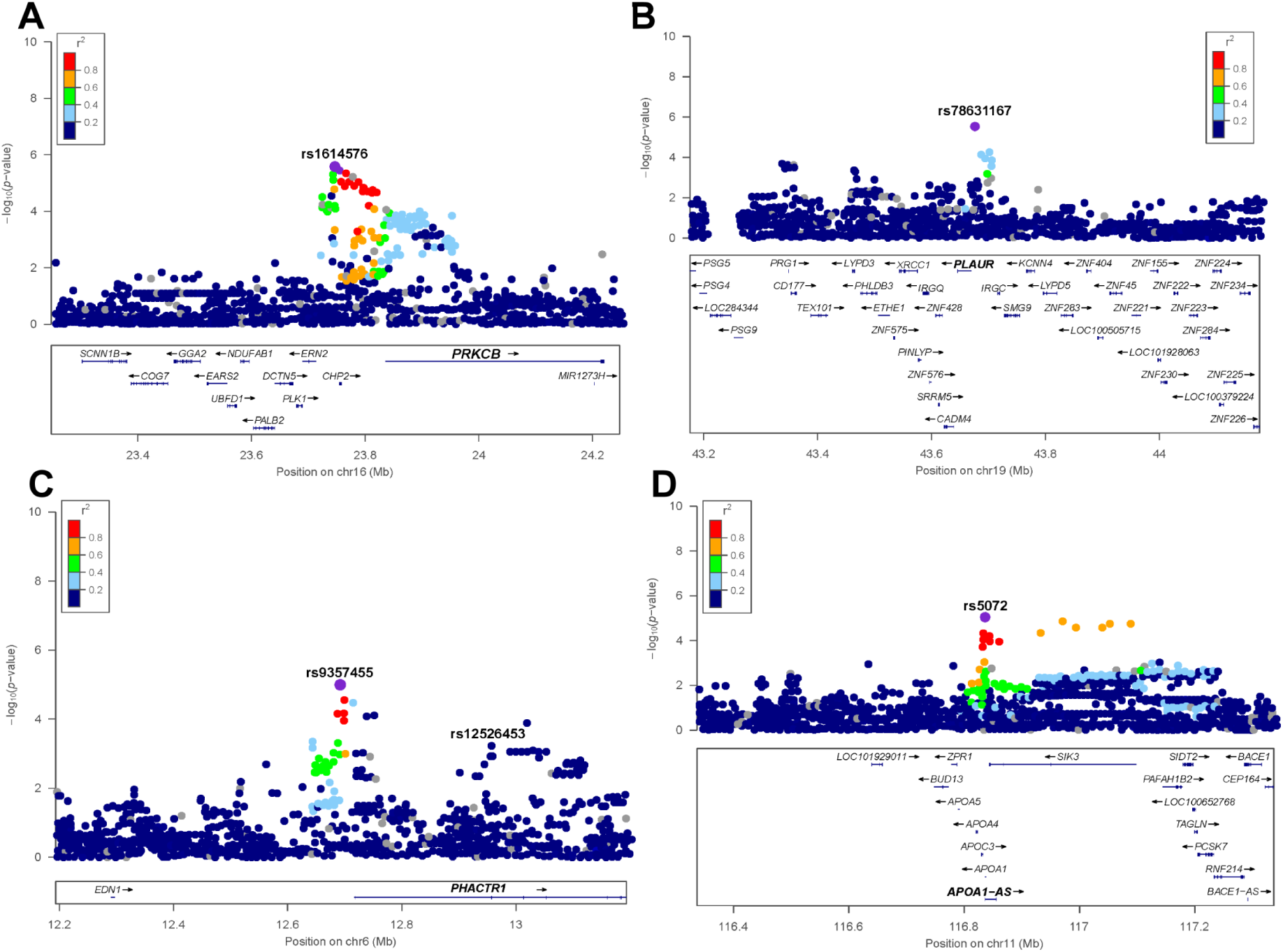
Regional association plot of genomic loci associated with early-onset AMI related to thrombosis, fibrinolysis, inflammation, and lipid metabolism. (A-D) Regional association plot at (A) rs1614576 (B) rs78631167 (C) rs9357455 (D) rs5072. For each plot, the x-axis indicates the genomic positions (hg38). Y-axis indicates -log_10_ *P*-value from the GWAS.

### Quantitative Trait Loci Mapping of the Novel Loci

Fourteen of the twenty-nine lead variants tended to have larger effect sizes in this study compared to the European (UK biobank; http://www.nealelab.is/uk-biobank/) and Japanese MI (Biobank of Japan) studies (20) (Table 2 and Supplementary Material, Fig. S4 and S5). The rest of the lead variants were not reported in previous SNP chip-based studies, implying that only limited genomic regions had been analyzed (22) and that WGS-based GWAS can identify novel loci using wider coverage of target genomic loci compared to SNP chip-based GWAS. Among the 29 novel loci for early-onset AMI, five loci appeared as eQTL based on the GTEx database (Fig. 3A and Supplementary Material, Fig. S6) (23, 24). Rs1614576 located upstream of the *PRKCB* had the most significant eQTL signals in whole blood (eQTL Bonferroni adjusted *P* = 4 × 10^−207^) by the eQTLGen database and artery tibial (*P* = 2.75 × 10^−14^; Fig. 3B) and artery aorta (*P* = 1.22 × 10^−9^) in the GTEx database (Supplementary Material, Table S8). The *PRKCB* gene affected by the rs1614576 loci is known to be involved in platelet responses implicated in thrombus formation (15). Rs591012 located in *TTK* gene was not only an eQTL (*P* = 4.34 × 10^−17^ in artery tibial; Fig. 3C), which affects the expression of the *ELOVL4* (ELOVL fatty acid elongase 4), involved in lipid metabolism, but also an sQTL that changes the alternative splicing of the gene (Supplementary Material, Table S8 and S9). Rs5072, located in *APOA1-AS*, exhibited eQTL signals that changed the expression of *PAFAH1B2* (platelet activating factor acetylhydrolase 1b catalytic subunit 2) (*P* = 1.68 × 10^−4^; Fig. 3D) and *PCSK7* (proprotein convertase subtilisin/kexin type 7) (*P* = 3.96 × 10^−11^) in cells cultured fibroblasts in the GTEx database (Supplementary Material, Table S8). In particular, *PAFAH1B2* encodes a subunit of PAFAH that degrades and inactivates platelet-activating factor (PAF), a potent platelet agonist (25). In addition, rs10232190 affects the expression of *LINC01003* (*P* = 1.00 × 10^−6^ and 1.16 × 10^−6^ in the heart atrial appendage and whole blood, respectively; Fig. 3E and Supplementary Material, Table S8). *LINC01003* can alter the expression of *PIM1* (proto-oncogene serine/threonine-protein kinase Pim-1) whose inhibition suppresses vascular smooth muscle cell (VSMC) proliferation and the development of atherosclerosis (26, 27).

**Figure 3.**
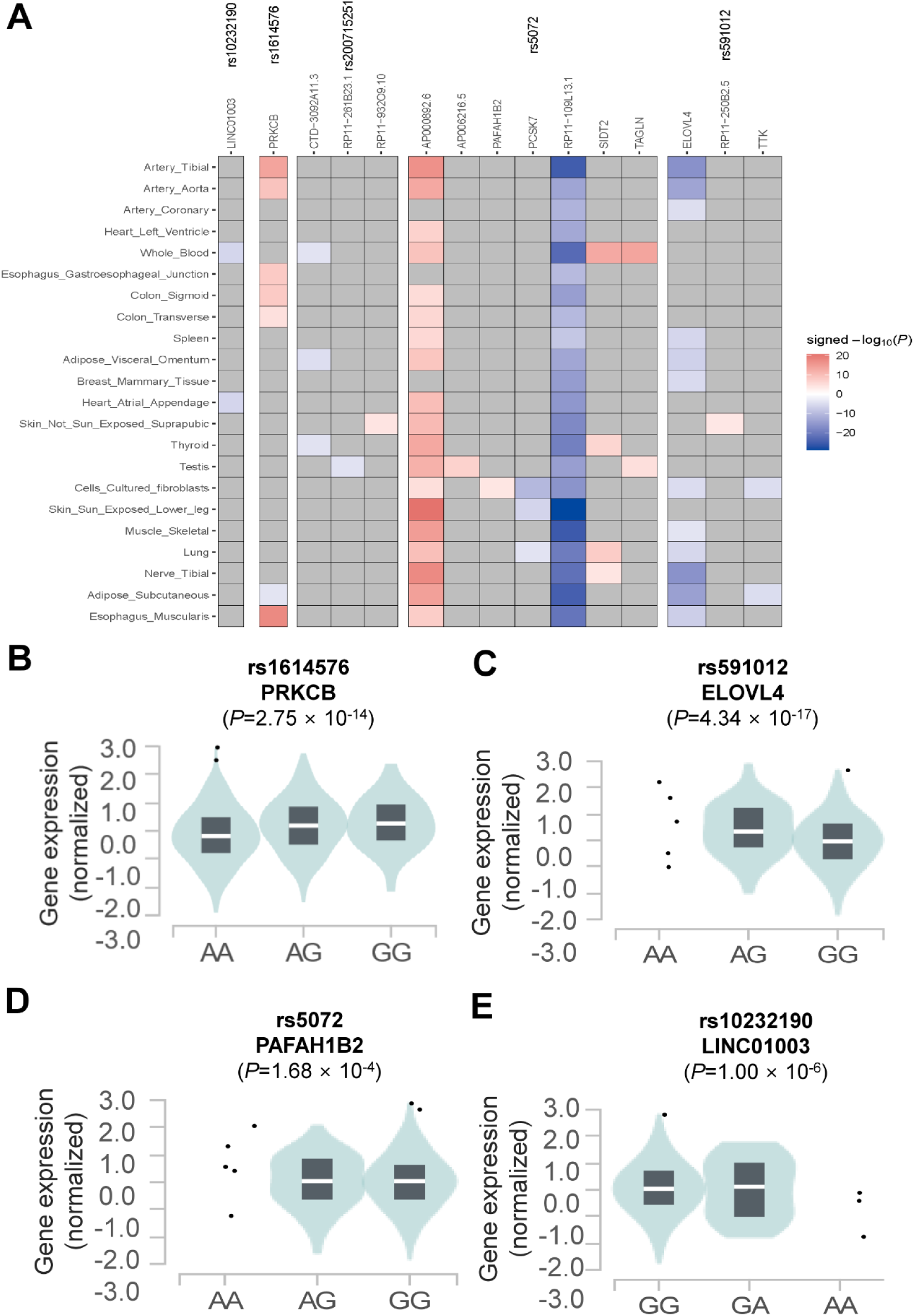
eQTL for the novel loci associated with early-onset AMI. (A) Heatmap of eQTL mapping to the novel loci for early-onset AMI. Rows show the tissue. Columns show the novel loci and the genes affected by the loci. Colors represent the signed - log_10_ *P*-value from the eQTL results. (B-E) Violin plot of eQTL at (B) rs1614576 (C) rs591012 (D) rs5072 (E) rs10232190. The lower and upper border of the box correspond to the first and third quartiles, respectively, the central line indicates the median.

## DISCUSSION

This study aimed to identify new genomic loci associated with early-onset AMI and determine their functional mechanisms using GWAS, QTL mapping, and protein-protein interaction analyses. The results revealed 29 novel loci associated with early-onset AMI, of which two loci are related to thrombosis and fibrinolysis, and confirmed the role of inflammation and lipid metabolism in the development of early-onset AMI.

Consistent with a previous study showing that lipid metabolism-associated genes such as *LDLR* and *PCSK9* are critical for early-onset AMI (2), the present study also identified a novel lipid metabolism associated locus in *APOA1-AS* that regulates the expression of *APOA1* gene which encodes a component of HDL cholesterol and protects against atherosclerosis (21). The eQTL analysis based on the GTEx dataset showed that the expression of the *ELOVL4* (ELOVL fatty acid elongase 4) gene, which is involved in lipid metabolism, was affected by the novel early-onset AMI risk loci identified in this study. *ELOVL4* encodes a crucial enzyme that plays a role in the production of very-long-chain fatty acids (VLC-SFA and VLC-PUFA) (28).

Few genetic markers associated with thrombus formation and fibrinolysis in AMI have been discovered (5, 13). In 2021, Hartiala *et al*. discovered a thrombosis associated locus located in the *SLC44A3* (Solute Carrier Family 44 Member 3) gene using GWAS for AMI (5). XU *et al*. suggested that fibrinolysis related genes that encodes urokinase-type plasminogen activator (uPA) which might influence the susceptibility to AMI; however, the study did not scan whole-genome (13). Additionally, several studies have reported loci that are mainly associated with lipid and cholesterol metabolism involving in plaque development and progression (6, 8, 29); however, the results of the present study reveal novel loci related to thrombosis and fibrinolysis, which are the leading cause of clinical manifestation of AMI. This result was supported by the PPI analysis of the 29 novel loci, showing that the interacting genes belong to fibrinolysis related pathways, such as dissolution of fibrin clot (HSA-75205) and complement and coagulation cascades (hsa04610) (Supplementary Material, Table S5-7). Previous clinical studies have focused mostly on the role of thrombosis in AMI formation (30). The spontaneous lysis of a coronary thrombus, which is a natural protective mechanism against lasting occlusion and myocardial infarction, is an important mechanism in the development of AMI (31). Although arterial thrombosis is responsible for most AMI cases, endogenous fibrinolysis is also a powerful defense mechanism against lasting arterial thrombotic occlusion (12). Recent clinical studies have reported that impaired endogenous fibrinolysis can be detected in a significant number of patients with the acute coronary syndrome, and this appears to be a novel risk factor that is independent of traditional cardiovascular risk factors and unaffected by antiplatelet therapy (12, 16, 32-34). Since only patients with early-onset AMI with definite AMI phenotypes were included in the study, the present study implies that patients with early-onset AMI may have susceptible genomic loci in the thrombosis related *PRKCB* gene and protective loci in the fibrinolysis related *PLAUR* gene, suggesting the association of these mechanisms with early-onset AMI.

Our study had three key strengths: clearly defined sample collection, whole-genome coverage, and strict removal of batch effects. This study collected early-onset AMI samples that underwent PCI for atherothrombotic occlusive lesions to prevent misclassification of AMI phenotype. Notably, AMI samples with non-obstructive coronary arteries were excluded to identify the cause of atherothrombosis, which has allowed us to acquire a considerable number of newly detected loci in our GWAS, despite the sample size. Additionally, previous studies on AMI utilized SNP chips such as Illumina human exome beadchip, Humanhap 610 Quad Chip, or Affymetrix 6.0 GeneChip (2, 5, 14) and only limited genomic regions could be covered by the SNP chip-based experiments (22). In 2019, Amit *et al*. studied the whole-genome of 2,081 patients with early-onset AMI; however, their study focused on analyzing the polygenic risk score rather than determining early-onset AMI-specific markers (35). In the present study, WGS data were analyzed to characterize the entire genome to identify early-onset AMI-specific markers, revealing 86% loci and 77% variants associated with early AMI that were not covered in SNP chips (Supplementary Material, Table S2 and S3). Moreover, the variants were strictly filtered to remove false positives caused by the batch effect which can lead to false disease association (36, 37). A batch effect was confirmed according to the sequencing year, and it was not successfully filtered by common variant filtering criteria such as missing genotype rate, minor allele frequency, and Hardy-Weinberg equilibrium (Supplementary Material, Fig. S7). The batch effect was removed by filtering the variants with allele balance bias, which identified systematic genotyping errors (36) (Supplementary Material, Fig. S8 and S9).

This study had two potential limitations. First, the sample size of this study was only one-third of that in the previous study by Amit *et al*. (35); however, our samples were clinically approved samples requiring PCI for atherothrombotic AMI, and our genetic analysis covered the whole genome. Second, we analyzed early-onset AMI in the Korean population. Therefore, the results need to be validated in multiracial populations. Nevertheless, the genomic loci identified in this study have four critical functions: thrombosis, fibrinolysis, inflammation, and lipid metabolism, which are known to be related to AMI. Therefore, these loci are generally applicable to other ethnic groups. We aim to verify the candidate loci using additional samples in functional studies with additional multi-omics data, including RNA and protein expression levels and epigenetic markers. These multi-layer omics data can provide researchers with detailed and conclusive understanding on AMI development.

In conclusion, this study identified 29 novel loci that are involved in all the steps of AMI development, including thrombosis and fibrinolysis, as well as previously known metabolic pathways, such as inflammation and lipid metabolism, using WGS data accompanied by GWAS, PPI, and QTL mapping. These results provide novel insights into the etiology of AMI. These novel genetic markers are potential biomarkers for the prediction, prevention, and personalized treatment of AMI in individuals who carry the loci.

## MATERIALS AND METHODS

### Data Sources and Study Population

In this study, 596 patients with early-onset AMI from four teaching hospitals in Korea were enrolled and 643 healthy subjects from the Korean Genome Project (KGP) were selected as control (38, 39). Patients with early-onset AMI were hospitalized with a diagnosis and treatment of an ST-segment elevation myocardial infarction (STEMI) or non-ST-segment elevation myocardial infarction (NSTEMI) caused by atherothrombotic occlusive lesions which were treated with PCI (40). Patients with early-onset AMI included in this study were men and women under 50 and 60 years of age, respectively. Population-based control subjects were derived from the KGP (38, 39). KGP is the largest Korean Genome Project and currently includes about 4,000 human genomes sequenced in Korea. Information regarding the KGP data can be found on the Korean Genome Project web page (http://koreangenome.org). The genomes of these 596 patients were compared with those of 643 control subjects in Korea, who were free of cardiovascular disease, between 2018 and 2019. All study participants provided written informed consent. Sample collection and sequencing were approved by the institutional review board (IRB) of the Ulsan National Institute of Science and Technology (UNISTIRB-15-19-A). Analyses were performed using Python version 3.7.7 and R version 3.5.0 (41, 42).

### Whole-genome sequencing by Illumina NovaSeq sequencer

WGS was performed using the Illumina NovaSeq 6000 platform and clinical information from the KGP was matched (38, 39). Of the 596 patient samples, 265 had already been sequenced in a previous study (40), and the remaining 331 samples were sequenced for this study (Table 1). Genomic DNA was isolated from the plate using a DNeasy Blood & Tissue kit (Qiagen, Germany) according to the manufacturer’s protocol. The extracted DNA was quantified using a Quant-iT BR assay kit (Invitrogen, USA). High–molecular-weight genomic DNA was sheared using a Covaris S2 ultrasonicator system to obtain fragments of appropriate sizes. Libraries with short 350–base pair (bp) inserts for paired-end reads were prepared using the TruSeq Nano DNA sample prep kit in accordance with the manufacturer’s protocol for Illumina-based sequencing. The products were quantified using a Bioanalyzer 2100 (Agilent, Santa Clara, CA, USA) and raw data were generated using an Illumina NovaSeq 6000 platform (Illumina). Clusters were generated using paired-end 2 × 150-bp cycle sequencing reads via resequencing. The quality of the bases in the reads was checked using FastQC (ver. 0.11.5; www.bioinformatics.babraham.ac.uk/projects/fastqc/) software. Additional details regarding genomic variant identification are provided in the Supplementary Methods in the Supplementary Material.

### Genome-Wide Association Study (GWAS)

GWAS was performed using logistic regression with an additive genetic model using PLINK (ver. 1.9b) (43). Additional details regarding batch effect removal and individual and variant filtering are provided in the Supplementary Methods in the Supplement Material. A total of 7,869,081 SNPs and indels were tested. Sex and the top ten principal components were included in the model as covariates. The genome-wide suggestive threshold was determined to be 1 × 10^−5^ for suggestive variants. Suggestive variants were considered novel according to the following criteria: (i) single nucleotide variants (SNVs) with no overlap with previously reported loci for myocardial infarction and (ii) SNVs with lead variants that do not have LD variants (*r*^*2*^ > 0.1) in previously reported loci for myocardial infarction. Variants were grouped into the loci with the ‘--clump-r2 0.1 –clump-window-size 250’ option with PLINK (ver. 1.9b) (43).

### Quantitative Trait Loci (QTL) Mapping

Variants were mapped to expression QTL (eQTL), splicing QTL (sQTL), and methylation QTL (mQTL) using publicly available datasets. For eQTL, which explains a fraction of the genetic variance of a gene expression phenotype (44), we mapped the lead variants from the GWAS to the eQTL from GTEx and eQTLGen dataset (23, 24). For sQTL, sQTL was determined using the GTEx dataset (23). For mQTL, mQTLdb, a large-scale genome-wide DNA methylation analysis of 1,000 mother-child pairs at series time points across the life-course (45), was used. QTLs for suggestive lead variants were reported.

## Data Availability

Full summary statistics relating to the GWAS analysis and other relevant data are available upon request to the authors.

## DATA AVAILABILITY

Full summary statistics relating to the GWAS analysis will be deposited on the Korean Genome Project web page (http://koreangenome.org/Cardiomics). All other relevant data are available upon request from the authors.

## ACKNOWLEDGEMENTS

We thank all the genome donating participants. The biospecimens for this study were provided by Ulsan Medical Center and the Biobanks of Gyeongsang National University Hospital, Chungbuk National University Hospital (18-27, 20-04), and Kyung Hee University Hospital (2018-4, 2019-4, 2019-6), the members of the National Biobank of Korea, which is supported by the Ministry of Health, Welfare and Family Affairs. We thank the Korea Institute of Science and Technology Information (KISTI) which provided us with the Korea Research Environment Open NETwork (KREONET).

The Genotype-Tissue Expression (GTEx) Project was supported by the Common Fund of the Office of the Director of the National Institutes of Health, and by NCI, NHGRI, NHLBI, NIDA, NIMH, and NINDS. The data used for the analyses described in this manuscript were obtained from the GTEx Portal on 11/25/2021.

## ABBREVIATIONS

AMI: acute myocardial infarction
PCI: percutaneous coronary intervention
GWAS: genome-wide association study
WGS: whole-genome sequencing
SNV: single nucleotide variant
KGP: Korean Genome Project
STEMI: ST-segment elevation myocardial infarction
NSTEMI: non-ST-segment elevation myocardial infarction
QTL: quantitative trait loci
eQTL: expression QTL
sQTL: splicing QTL
mQTL: methylation QTL
PRKCB: protein kinase C beta
PLAUR: plasminogen activator, urokinase receptor
PAFAH1B2: platelet-activating factor acetylhydrolase 1b catalytic subunit 2
PHACTR1: phosphatase and actin regulator 1
APOA1-AS: apolipoprotein A1 antisense RNA
PPI: protein-protein interaction

