## Supplementary Material for "Genome-wide analyses of early-onset acute myocardial infarction identify 29 novel loci by whole genome sequencing"

### SUPPLEMENTAL MATERIAL

### Supplemental Methods

#### Genomic variants identification

We trimmed the adapter contamination of the sequencing reads using Cutadapt (ver. 1.9.1) with a forward adapter (′GATCGGAAGAGCACACGTCTGAACTCCAGTCAC′) and reverse adapter (′GATCGGAAGAGCGTCGTGTAGGGAAAGAGTGT′) and with a minimum read length of 50 bp after trimming.[1] Then, the trimmed reads were mapped to the human genome reference, hg38 using BWA-MEM (ver. 0.7.16a) with the "-M" option and alt-aware mode.[2] We sorted the mapped BAM files by coordination using Picard ("Picard Toolkit." 2019. Broad Institute, GitHub Repository. https://broadinstitute.github.io/picard/; Broad Institute) (ver. 2. 14. 0) with the Sortsam module. Duplicated reads were marked using Picard (ver. 2.14.0) with the MarkDuplicates module. We recalibrated the mapping quality using BaseRecalibrator tool in the Genome Analysis Tool Kit (GATK) (ver. 4.1.3).[3] gVCF files of the individuals were generated by HaplotypeCaller in GATK with the "--genotyping-mode DISCOVERY -stand-call-conf 30 -ERC GVCF" option.[3] We merged gVCF files of all individuals per chromosome using CombineGVCFs in GATK. SNVs and indels were jointly genotyped from the merged gVCF files by GenotypeGVCFs in GATK. We annotated variant qualities by VariantQualityScoreRecalibration (VQSR) in GATK. All variants were annotated using the Ensembl VEP (ver. 92.1) and ANNOVAR.[4, 5]

#### Batch effect removal

Each sample was labeled in accordance with its sequencing year. Two technical batches were identified. The batch effect was assessed via PCA, using PLINK (ver. 1.9b),[6] using variants in accordance with the following criteria (Figure S3-5):

1. Biallelic SNVs with a MAF of ≥ 1%
2. *P* values of the Hardy-Weinberg Equilibrium (HWE) test > 0.000001
3. Genotype missing rate of < 0.01.

Thereafter, the filtered variants were pruned on the basis of linkage disequilibrium (LD), using PLINK (ver. 1.9b) [6] with '--indep-pairwise 200 4 0.1' option.

We filtered out the variants according to the following criteria:

1. SNVs which fail Variant Quality Score Recalibration (VQSR)
2. Heterozygous SNVs which deviate ± standard deviation (SD) from their mean of allele balance (AB) were removed from the dataset.

Allele balance (AB) denotes the fraction of reads supporting the alternative allele in a focal position (AB = alternative read count/total read count at focal position).[7]

### Individual and variant filtering for GWAS

1,159 individuals and 7,869,081 variants were selected in accordance with the following criteria:

For individuals:

1. Individuals whose rate of genotype missingness is less than 10%
2. Individuals who don't deviate ± 3 SD from the samples' heterozygosity rate mean
3. Individuals who deviate ± 4 Z-score of identity-by-state (IBS) distance of 1-5th nearest neighbor
4. Individuals who have no kinship
5. Individuals who are not ancestry outliers

For variants:

1. SNVs and indels having bi-allele
2. SNVs and indels having a MAF ≥ 1%
3. SNVs and indels having an HWE $P > 10^{-6}$
4. SNVs and indels having a missing genotype rate of < 0.01

**Protein-Protein Interaction (PPI)**

Protein-Protein Interaction network was constructed by the suggestive loci associated with early-onset AMI. We utilized STRING database (ver. 11.5) that contains the information of interactions including both physical and functional associations.[8] Functional enrichment test was conducted in the PPI network using REACTOME and KEGG pathway database.

### Supplemental Figures

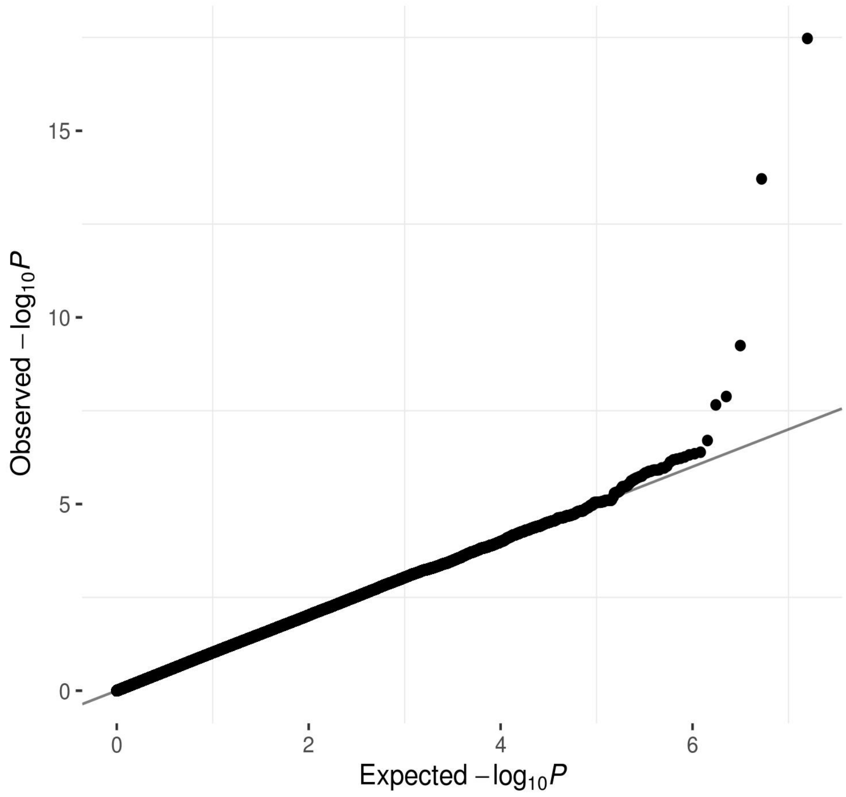


**Supplementary Figure 1. QQplot for the GWAS of the early-onset AMI.**

X-axis indicates expected -$\log_{10}$ *P*-value. Y-axis indicates observed -$\log_{10}$ *P*-value.

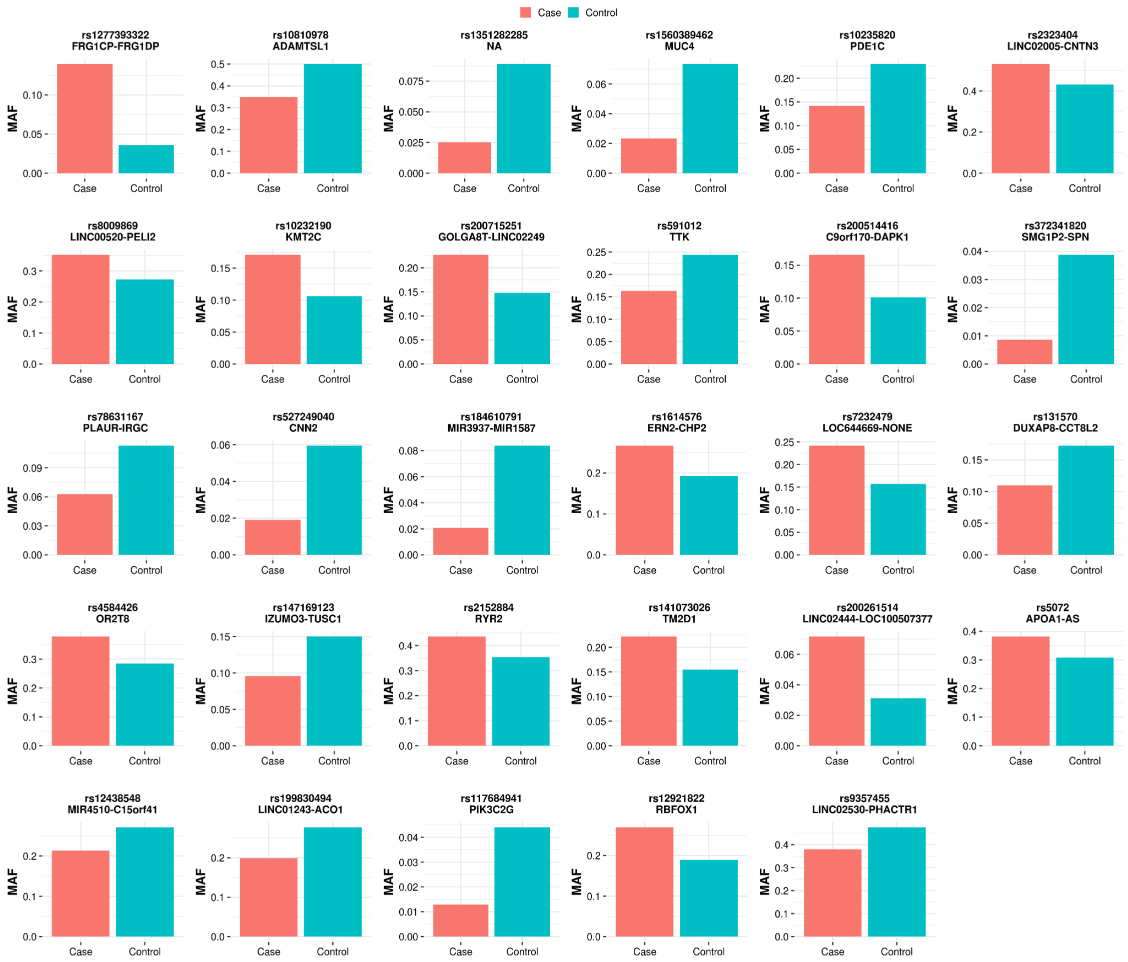


**Supplementary Figure 2. Minor allele frequency distribution for 29 suggestive loci between early-onset AMI and healthy subjects**

X-axis indicates the group. Case and control represent early-onset AMI and healthy subjects, respectively. Y-axis indicates minor allele frequency.

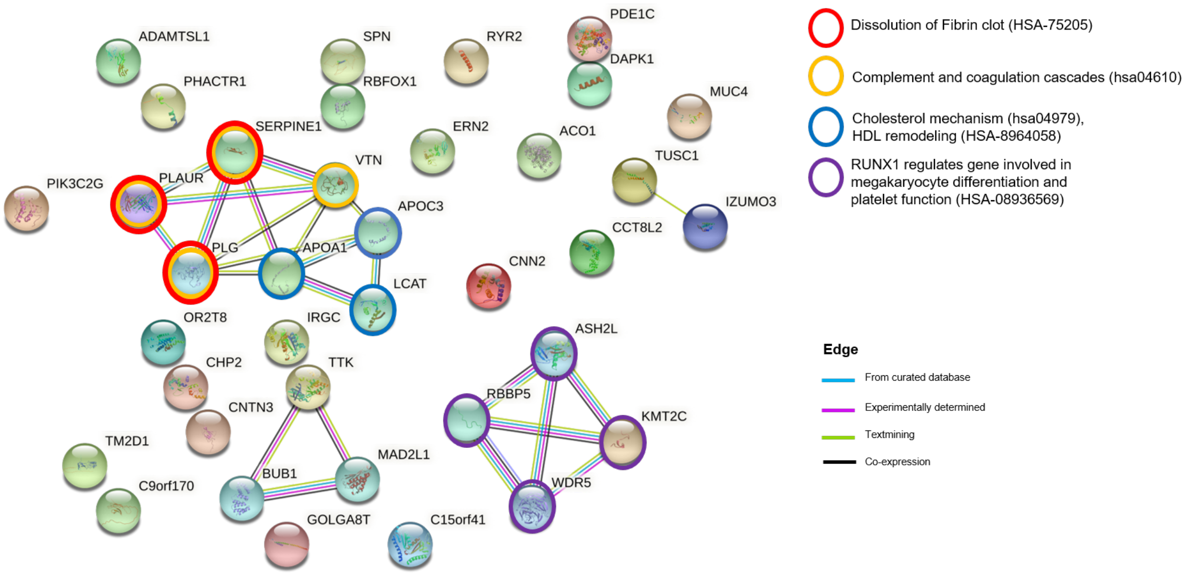


**Supplementary Figure 3. Protein-Protein Interaction (PPI) network of the suggestive loci**

PPI network plot of the suggestive loci associated with early-onset AMI. Edges denote protein-protein associations. Colors of circle represent biological pathways to which the protein belongs. PPI network is based on STRING database (ver. 11.5).[8]

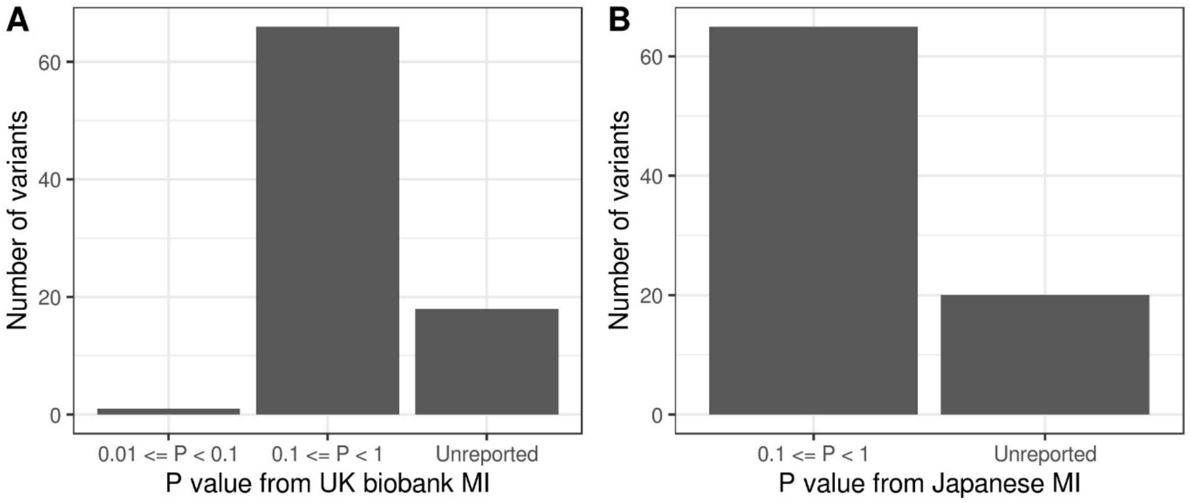


**Supplementary Figure 4. *P*-value distribution of 85 suggestive variants compared to GWAS results for myocardial infarction from Japanese and UK biobank**

X-axis indicates the *P*-value of GWAS results for myocardial infarction from (A) UK biobank and (B) biobank of Japan. Y-axis indicates the number of variants.

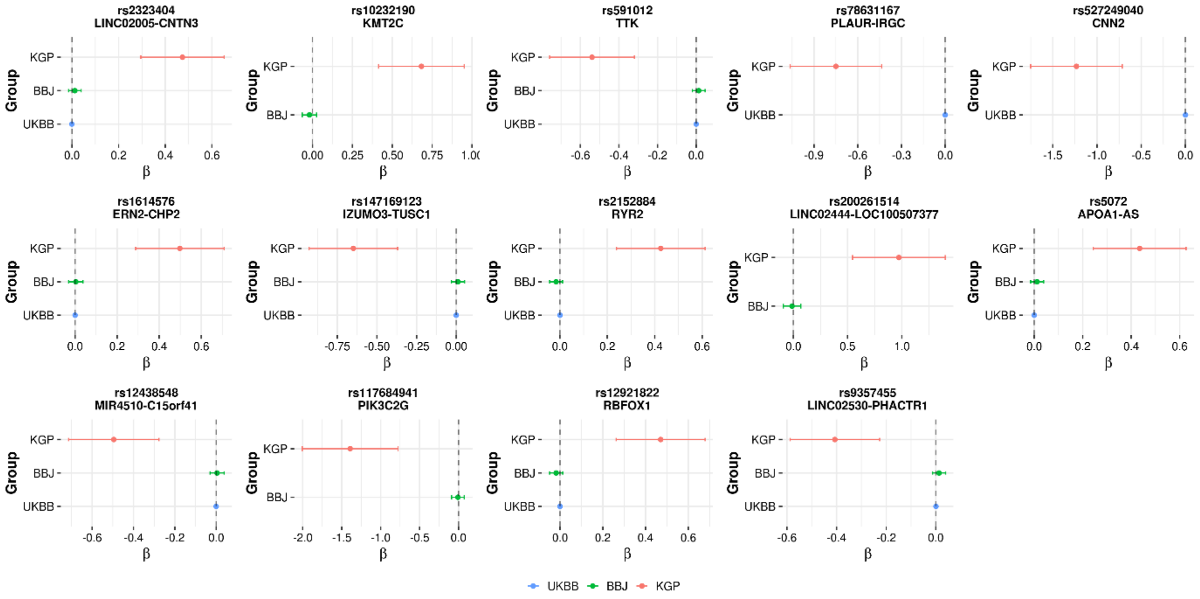


**Supplementary Figure 5. Comparison of effect size of the novel loci for early-onset AMI**

X-axis indicates the effect size (beta) of the novel loci. Error bar denotes 95% confidence interval (CI). Y-axis indicates the GWAS dataset. KGP, Korean Genome Project; BBJ, Biobank of Japan; UKBB, UK biobank.

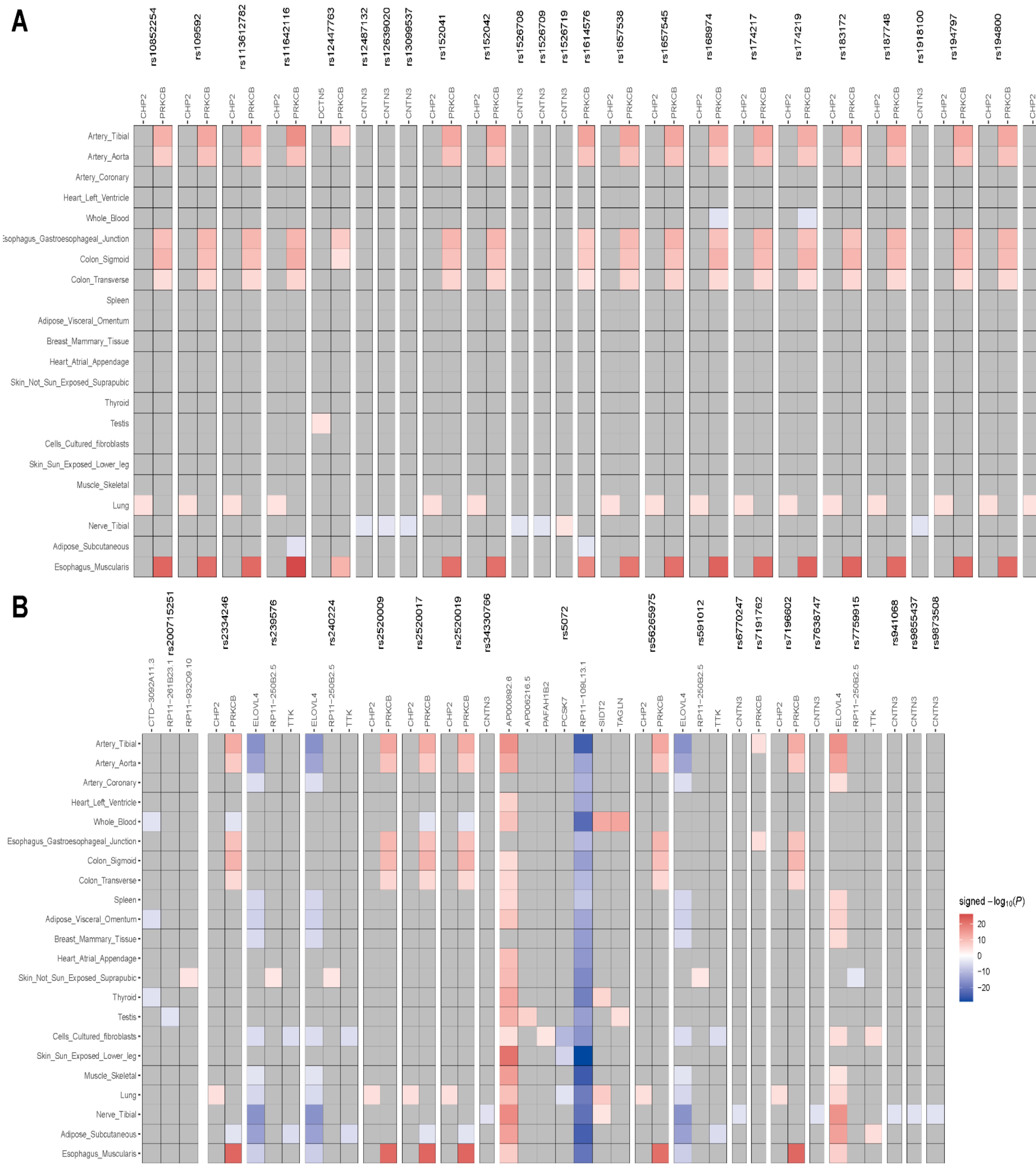


**Supplementary Figure 6. Heatmap of eQTL mapping to the suggestive variants for early-onset AMI**

Rows show the tissue. Columns show the novel loci and the genes affected by the loci. Colors represent the signed -log$_{10}$ *P*-value from the eQTL results.

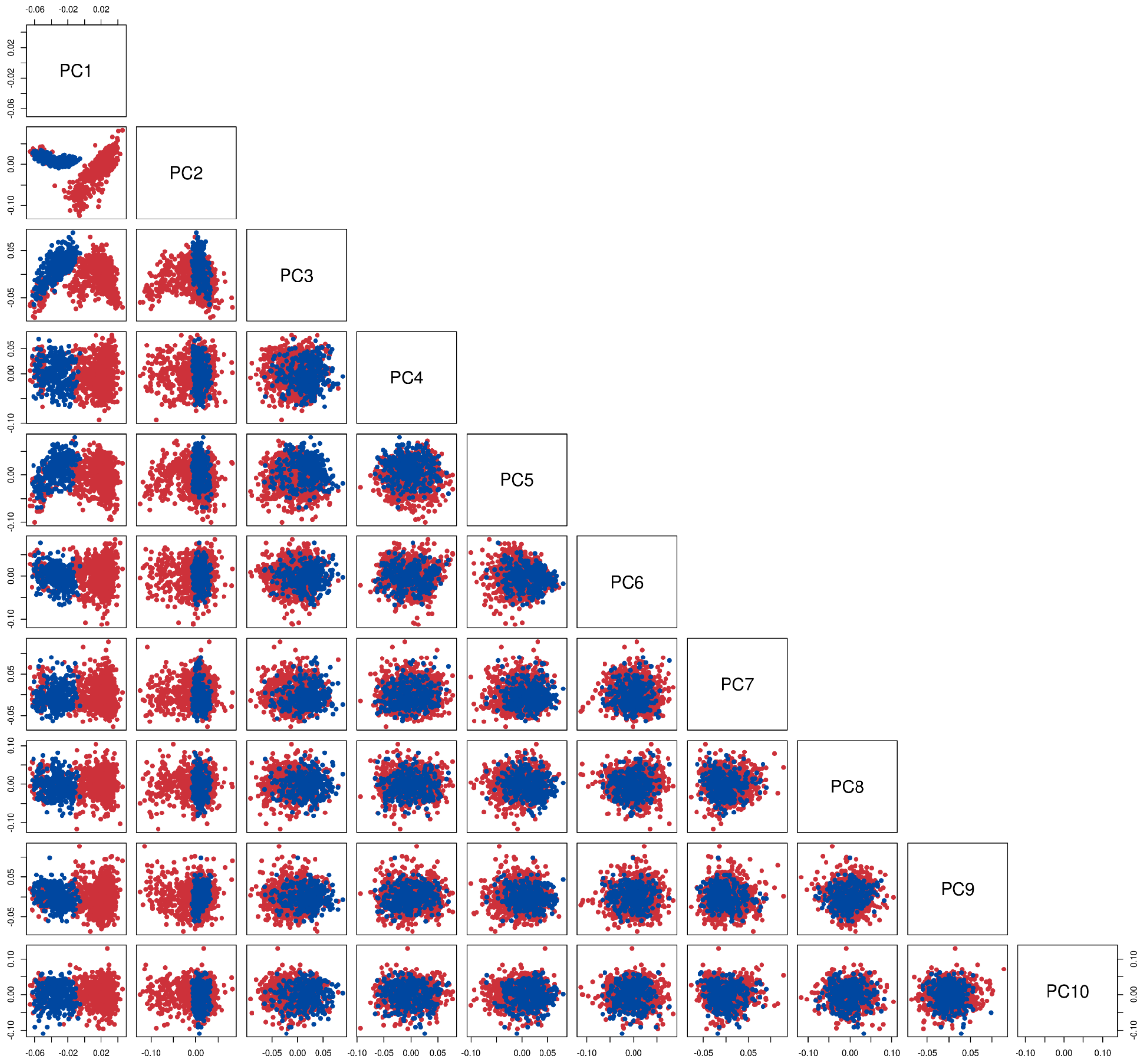


**Supplementary Figure 7. PCA using variants before VQSR and AB filtering**

Pairs plot of the top ten dimensions of the PCA. Colors denote technical batches. Red and blue indicate samples sequenced in 2018 and 2019, respectively.

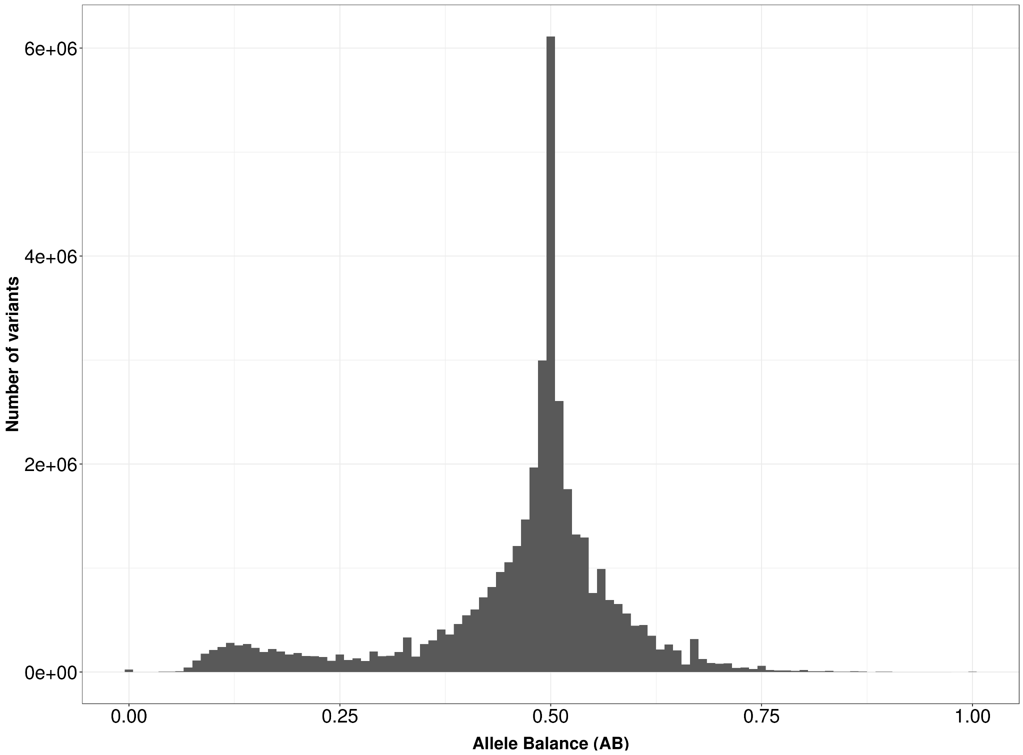


**Supplementary Figure 8. Allele balance distribution for heterozygous variants**

X-axis indicates allele balance (AB). It denotes the fraction of reads supporting the alternative allele in a focal position (AB = alternative read count/total read count at focal position) [7]. Y-axis indicates the number of variants.

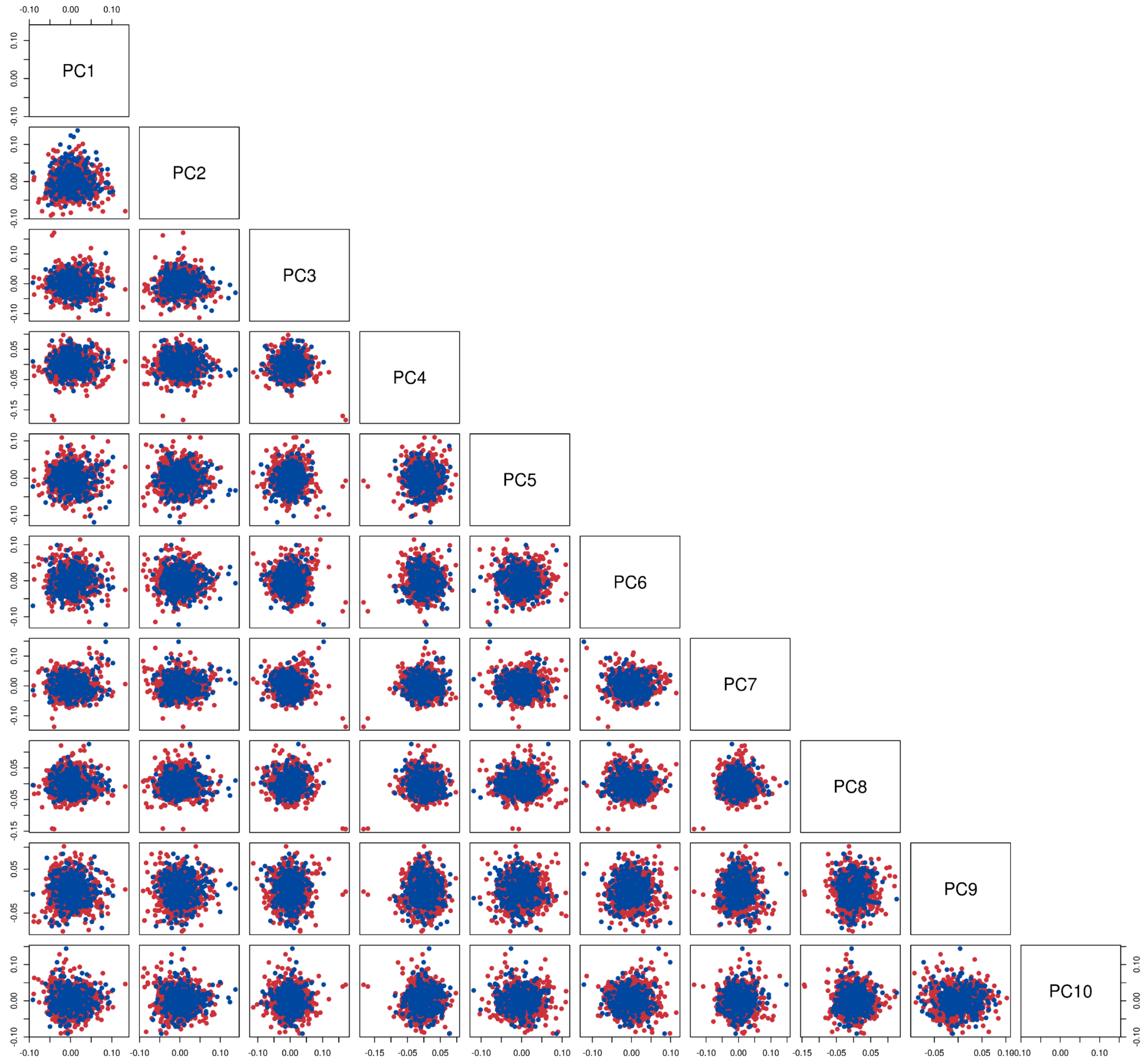


**Supplementary Figure 9. PCA using variants after VQSR and AB filtering**

Pairs plot of the top ten dimensions of the PCA. Colors denote technical batches. Red and blue indicate samples sequenced in 2018 and 2019, respectively.

### Supplemental Tables

#### Supplementary Table 1. Suggestive variants for early-onset AMI compared to European and Japanese MI studies

| Gene | Chromosome | Position | rs ID | REF | ALT | Effect allele | Korean Genome Project (early-onset AMI in Korean) | | BBJ (MI in Japanese) | | UK biobank (MI in European) | |
|---|---|---|---|---|---|---|---|---|---|---|---|---|
| | | | | | | | Odds ratio | *P* | BETA | *P* | BETA | *P* |
| FRG1CP-FRG1DP | 20 | 28,772,995 | rs1277393322 | G | A | A | 5.713 | 3.36E-18 | NA | NA | NA | NA |
| ADAMTSL1 | 9 | 18,523,845 | rs10810978 | C | A | C | 0.451 | 1.94E-14 | NA | NA | NA | NA |
| NA | 3 | 164,704,630 | rs1351282285 | TTAAATG | T | T | 0.239 | 5.67E-10 | NA | NA | NA | NA |
| MUC4 | 3 | 195,788,126 | rs1560389462 | GGGTGGTGTGACCTGTGGATACTGAGGAAGTGTCGGTGACAGGAAGAGAGGTGGCGTGACCTGTGGATGCTGAGGAAGTGTCGGTGACAGGAAGAGGGGTGGTGTGACCTGTGGATACTGAGGAAGTGTCGGTGACAGGAAGAGA | G | G | 0.256 | 1.31E-08 | NA | NA | NA | NA |
| PDE1C | 7 | 32,082,944 | rs10235820 | A | G | A | 0.497 | 2.21E-08 | NA | NA | NA | NA |
| LINC02005-CNTN3 | 3 | 73,926,364 | rs2323404 | A | G | A | 1.606 | 1.98E-07 | 0.012243 | 3.68E-01 | -0.000594 | 2.32E-01 |
| LINC02005-CNTN3 | 3 | 73,923,420 | rs7641082 | T | C | T | 1.585 | 4.08E-07 | 0.014099 | 2.99E-01 | -0.000572 | 2.49E-01 |
| LINC00520-PELI2 | 14 | 55,900,262 | rs8009869 | A | G | G | 1.653 | 4.49E-07 | NA | NA | NA | NA |
| LINC02005-CNTN3 | 3 | 73,921,969 | rs1405402 | G | A | G | 1.579 | 4.81E-07 | NA | NA | -0.000640 | 2.01E-01 |
| LINC02005-CNTN3 | 3 | 73,927,087 | rs1918101 | G | T | G | 1.576 | 5.47E-07 | 0.011962 | 3.80E-01 | -0.000582 | 2.47E-01 |
| LINC02005-CNTN3 | 3 | 73,923,383 | rs7641013 | T | C | T | 1.574 | 5.92E-07 | 0.014116 | 2.99E-01 | -0.000537 | 2.78E-01 |
| KMT2C | 7 | 152,231,944 | rs10232190 | G | A | A | 1.980 | 6.24E-07 | -0.019416 | 3.96E-01 | NA | NA |
| GOLGA8T-LINC02249 | 15 | 30,175,826 | rs200715251 | A | G | G | 1.840 | 6.56E-07 | NA | NA | NA | NA |
| LINC02005-CNTN3 | 3 | 73,927,219 | rs1918100 | T | A | T | 1.764 | 7.45E-07 | 0.004968 | 7.62E-01 | -0.000365 | 2.58E-01 |
| LINC02005-CNTN3 | 3 | 73,925,188 | rs9855437 | A | G | A | 1.754 | 9.60E-07 | 0.005191 | 7.52E-01 | -0.000395 | 2.20E-01 |
| LINC02005-CNTN3 | 3 | 73,925,641 | rs941068 | C | T | C | 1.748 | 1.08E-06 | 0.004697 | 7.75E-01 | -0.000392 | 2.24E-01 |

| | | | | | | | | | | | | |
|---|---|---|---|---|---|---|---|---|---|---|---|---|
| LINC02005-CNTN3 | 3 | 73,927,363 | rs6770247 | G | A | G | 1.748 | 1.08E-06 | 0.004966 | 7.62E-01 | -0.000358 | 2.67E-01 |
| LINC02005-CNTN3 | 3 | 73,927,320 | rs1526719 | A | G | G | 1.745 | 1.20E-06 | 0.003641 | 8.26E-01 | 0.000368 | 2.54E-01 |
| LINC00520-PELI2 | 14 | 55,906,845 | rs80334730 | T | C | C | 1.615 | 1.22E-06 | 0.010474 | 4.45E-01 | 0.000632 | 6.51E-01 |
| LINC00520-PELI2 | 14 | 55,907,084 | rs77800378 | G | A | A | 1.615 | 1.22E-06 | 0.010474 | 4.45E-01 | 0.000591 | 6.72E-01 |
| LINC02005-CNTN3 | 3 | 73,919,134 | rs12487132 | A | C | A | 1.747 | 1.26E-06 | 0.009700 | 5.54E-01 | -0.000347 | 2.82E-01 |
| LINC02005-CNTN3 | 3 | 73,925,782 | rs9873508 | G | A | G | 1.740 | 1.33E-06 | 0.004959 | 7.63E-01 | -0.000392 | 2.24E-01 |
| LINC02005-CNTN3 | 3 | 73,918,804 | rs1526708 | G | A | G | 1.744 | 1.34E-06 | 0.010012 | 5.40E-01 | -0.000349 | 2.79E-01 |
| LINC00520-PELI2 | 14 | 55,908,821 | rs1958821 | G | C | C | 1.614 | 1.44E-06 | 0.008646 | 5.28E-01 | 0.000581 | 6.78E-01 |
| TTK | 6 | 80,038,900 | rs591012 | G | A | G | 0.583 | 1.47E-06 | 0.013298 | 4.28E-01 | 0.000187 | 5.73E-01 |
| LINC02005-CNTN3 | 3 | 73,918,900 | rs1526709 | C | A | C | 1.736 | 1.59E-06 | 0.009565 | 5.59E-01 | -0.000344 | 2.88E-01 |
| C9orf170-DAPK1 | 9 | 87,414,203 | rs200514416 | T | C | C | 1.948 | 1.78E-06 | NA | NA | NA | NA |
| LINC00520-PELI2 | 14 | 55,905,697 | rs111432322 | C | T | T | 1.601 | 1.83E-06 | 0.010860 | 4.28E-01 | 0.000673 | 6.29E-01 |
| LINC02005-CNTN3 | 3 | 73,924,589 | rs34330766 | A | G | A | 1.731 | 1.88E-06 | 0.007019 | 6.71E-01 | -0.000386 | 2.31E-01 |
| LINC02005-CNTN3 | 3 | 73,923,365 | rs7638747 | A | G | A | 1.728 | 1.97E-06 | 0.008631 | 5.99E-01 | -0.000400 | 2.15E-01 |
| NA | 14 | 55,905,174 | rs10589173 | ATATAT | A | A | 1.598 | 2.02E-06 | 0.010768 | 4.32E-01 | 0.000456 | 7.49E-01 |
| LINC02005-CNTN3 | 3 | 73,919,588 | rs12639020 | C | T | C | 1.722 | 2.19E-06 | 0.009621 | 5.57E-01 | -0.000380 | 2.39E-01 |
| LINC02005-CNTN3 | 3 | 73,919,636 | rs12639023 | C | T | C | 1.722 | 2.19E-06 | NA | NA | -0.000375 | 2.45E-01 |
| TTK | 6 | 80,021,249 | rs239576 | G | A | G | 0.589 | 2.41E-06 | 0.013177 | 4.32E-01 | 0.000186 | 5.73E-01 |
| TTK-BCKDHB | 6 | 80,062,270 | rs7759915 | C | T | T | 0.589 | 2.41E-06 | 0.014708 | 3.81E-01 | -0.000193 | 5.59E-01 |
| SMG1P2-SPN | 16 | 29,655,837 | rs372341820 | G | A | A | 0.180 | 2.75E-06 | NA | NA | NA | NA |
| PLAUR-IRGC | 19 | 43,676,115 | rs78631167 | T | C | C | 0.473 | 2.93E-06 | NA | NA | 0.000327 | 8.02E-01 |
| CNN2 | 19 | 1,029,328 | rs527249040 | G | C | C | 0.292 | 3.20E-06 | NA | NA | 0.001111 | 7.53E-01 |
| MIR3937-MIR1587 | X | 39,788,017 | rs184610791 | C | G | G | 0.234 | 3.23E-06 | NA | NA | NA | NA |
| ERN2-CHP2 | 16 | 23,747,804 | rs1614576 | A | G | G | 1.646 | 3.41E-06 | 0.003445 | 8.44E-01 | 0.000026 | 9.44E-01 |
| ERN2-CHP2 | 16 | 23,747,877 | rs1657538 | G | A | A | 1.646 | 3.41E-06 | 0.003137 | 8.57E-01 | 0.000046 | 9.06E-01 |
| ERN2-CHP2 | 16 | 23,748,316 | rs1657545 | A | G | G | 1.646 | 3.41E-06 | 0.003101 | 8.59E-01 | 0.000056 | 8.86E-01 |
| TTK | 6 | 80,007,571 | rs240224 | G | T | G | 0.594 | 3.45E-06 | 0.013210 | 4.31E-01 | 0.000194 | 5.59E-01 |
| LOC644669-NONE | 18 | 15,377,667 | rs7232479 | G | A | A | 1.763 | 3.82E-06 | NA | NA | NA | NA |
| LINC02005-CNTN3 | 3 | 73,923,060 | rs13099537 | T | C | T | 1.696 | 4.17E-06 | 0.008677 | 5.97E-01 | -0.000398 | 2.17E-01 |
| CHP2 | 16 | 23,754,252 | rs152042 | A | G | G | 1.635 | 4.52E-06 | 0.004041 | 8.15E-01 | 0.000082 | 8.33E-01 |

| | | | | | | | | | | | | |
|---|---|---|---|---|---|---|---|---|---|---|---|---|
| DUXAP8-CCT8L2 | 22 | 16,400,343 | rs131570 | T | C | C | 0.544 | 4.66E-06 | NA | NA | NA | NA |
| ERN2-CHP2 | 16 | 23,741,456 | rs7191762 | G | A | A | 1.659 | 4.82E-06 | -0.003865 | 8.26E-01 | -0.000296 | 4.81E-01 |
| OR2T8 | 1 | 247,921,552 | rs4584426 | A | G | G | 1.538 | 4.83E-06 | NA | NA | NA | NA |
| CHP2 | 16 | 23,754,034 | rs152041 | G | C | C | 1.632 | 4.93E-06 | 0.003935 | 8.20E-01 | 0.000099 | 7.98E-01 |
| ERN2-CHP2 | 16 | 23,748,912 | rs11649332 | C | T | T | 1.632 | 4.98E-06 | NA | NA | 0.000090 | 8.17E-01 |
| IZUMO3-TUSC1 | 9 | 25,189,022 | rs147169123 | A | AT | A | 0.523 | 5.14E-06 | 0.011244 | 5.95E-01 | -0.000003 | 9.95E-01 |
| NA | 16 | 23,771,655 | rs55819522 | TAA | T | T | 1.624 | 6.41E-06 | 0.005604 | 7.44E-01 | 0.000087 | 8.22E-01 |
| LINC00520-PELI2 | 14 | 55,904,081 | rs6573071 | G | A | A | 1.531 | 6.72E-06 | 0.009769 | 4.70E-01 | 0.000646 | 6.44E-01 |
| ERN2-CHP2 | 16 | 23,742,273 | rs12447763 | G | T | T | 1.661 | 7.60E-06 | 0.003303 | 8.54E-01 | -0.000251 | 5.39E-01 |
| CHP2-PRKCB | 16 | 23,784,944 | rs2520017 | G | A | A | 1.616 | 7.94E-06 | 0.007308 | 6.71E-01 | 0.000095 | 8.06E-01 |
| CHP2-PRKCB | 16 | 23,785,567 | rs2520019 | T | C | C | 1.616 | 7.94E-06 | 0.007296 | 6.71E-01 | 0.000101 | 7.94E-01 |
| CHP2-PRKCB | 16 | 23,786,091 | rs11642116 | A | C | C | 1.616 | 7.94E-06 | 0.007194 | 6.76E-01 | 0.000055 | 8.88E-01 |
| CHP2-PRKCB | 16 | 23,786,294 | rs7196602 | C | T | T | 1.616 | 7.94E-06 | 0.007304 | 6.71E-01 | 0.000109 | 7.77E-01 |
| CHP2-PRKCB | 16 | 23,787,611 | rs2334246 | A | T | T | 1.616 | 7.94E-06 | 0.007250 | 6.73E-01 | 0.000107 | 7.82E-01 |
| CHP2-PRKCB | 16 | 23,788,500 | rs194817 | G | A | A | 1.616 | 7.94E-06 | 0.007240 | 6.74E-01 | 0.000107 | 7.82E-01 |
| CHP2-PRKCB | 16 | 23,789,042 | rs168974 | T | A | A | 1.616 | 7.94E-06 | 0.007150 | 6.78E-01 | 0.000108 | 7.80E-01 |
| CHP2-PRKCB | 16 | 23,789,509 | rs183172 | T | C | C | 1.616 | 7.94E-06 | 0.007168 | 6.77E-01 | 0.000118 | 7.61E-01 |
| RYR2 | 1 | 237,214,855 | rs2152884 | G | A | A | 1.530 | 8.02E-06 | -0.015869 | 2.55E-01 | 0.000728 | 4.58E-02 |
| TM2D1 | 1 | 61,717,206 | rs141073026 | A | C | C | 1.673 | 8.13E-06 | NA | NA | NA | NA |
| LINC02444-LOC100507377 | 12 | 73,222,956 | rs200261514 | G | GTAAT | GTAAT | 2.644 | 8.25E-06 | -0.011721 | 7.76E-01 | NA | NA |
| APOA1-AS | 11 | 116,836,867 | rs5072 | A | G | A | 1.545 | 8.61E-06 | 0.011949 | 3.82E-01 | -0.000046 | 9.41E-01 |
| CHP2 | 16 | 23,757,389 | rs109592 | C | T | T | 1.609 | 8.63E-06 | 0.004518 | 7.93E-01 | 0.000087 | 8.23E-01 |
| CHP2 | 16 | 23,759,927 | rs56265975 | T | G | G | 1.609 | 8.63E-06 | 0.005109 | 7.66E-01 | 0.000082 | 8.32E-01 |
| MIR4510-C15orf41 | 15 | 36,119,067 | rs12438548 | G | A | G | 0.609 | 8.72E-06 | 0.004081 | 8.14E-01 | -0.000343 | 3.30E-01 |
| LINC01243-ACO1 | 9 | 32,154,607 | rs199830494 | G | A | G | 0.631 | 8.77E-06 | NA | NA | NA | NA |
| CHP2-PRKCB | 16 | 23,761,303 | rs113612782 | G | A | A | 1.610 | 8.92E-06 | 0.005446 | 7.51E-01 | 0.000076 | 8.45E-01 |
| CHP2-PRKCB | 16 | 23,767,186 | rs187748 | G | A | A | 1.610 | 8.92E-06 | 0.005659 | 7.40E-01 | 0.000089 | 8.17E-01 |
| CHP2-PRKCB | 16 | 23,768,995 | rs174217 | A | G | G | 1.610 | 8.92E-06 | 0.005719 | 7.38E-01 | 0.000087 | 8.22E-01 |
| CHP2-PRKCB | 16 | 23,769,092 | rs2520009 | C | T | T | 1.610 | 8.92E-06 | 0.005791 | 7.35E-01 | 0.000090 | 8.17E-01 |
| CHP2-PRKCB | 16 | 23,769,518 | rs194797 | T | C | C | 1.610 | 8.92E-06 | 0.005732 | 7.37E-01 | 0.000087 | 8.22E-01 |
| PIK3C2G | 12 | 18,556,791 | rs117684941 | T | C | C | 0.249 | 8.95E-06 | -0.008694 | 8.35E-01 | NA | NA |
| LINC00520-PELI2 | 14 | 55,901,317 | rs4901620 | G | A | A | 1.523 | 8.96E-06 | 0.009618 | 4.76E-01 | 0.000396 | 5.50E-01 |

| | | | | | | | | | | | | |
|---|---|---|---|---|---|---|---|---|---|---|---|---|
| RBFOX1 | 16 | 7,002,773 | rs12921822 | C | T | C | 1.601 | 9.05E-06 | -0.018229 | 2.43E-01 | -0.000056 | 8.63E-01 |
| CHP2-PRKCB | 16 | 23,763,247 | rs194800 | T | C | C | 1.611 | 9.09E-06 | 0.007239 | 6.72E-01 | 0.000082 | 8.31E-01 |
| CHP2-PRKCB | 16 | 23,790,378 | rs10852254 | T | C | C | 1.605 | 9.12E-06 | 0.007414 | 6.66E-01 | 0.000125 | 7.48E-01 |
| CHP2-PRKCB | 16 | 23,790,834 | rs194815 | A | G | G | 1.605 | 9.12E-06 | 0.007345 | 6.69E-01 | 0.000132 | 7.34E-01 |
| CHP2-PRKCB | 16 | 23,791,390 | rs194813 | T | C | C | 1.605 | 9.12E-06 | 0.007373 | 6.67E-01 | 0.000139 | 7.19E-01 |
| CHP2-PRKCB | 16 | 23,791,599 | rs174219 | T | C | C | 1.605 | 9.12E-06 | 0.007367 | 6.68E-01 | 0.000144 | 7.11E-01 |
| LINC02530-PHACTR1 | 6 | 12,693,654 | rs9357455 | C | T | C | 0.666 | 9.73E-06 | 0.012942 | 3.34E-01 | 0.000479 | 3.12E-01 |

**Supplementary Table 2. List of variants covered by SNP chip among 29 novel genomic loci**

| rsID | Gene | SNP_chip_name | Probe_ID |
|---|---|---|---|
| rs591012 | TTK | Human_610_Quad | rs591012-131_T_F_1864142653 |
| rs2152884 | RYR2 | Affymetrix | SNP_A-8604630 |
| rs12921822 | RBFOX1 | Affymetrix | SNP_A-1782059 |
| rs9357455 | LINC02530-PHACTR1 | Human_610_Quad | rs9357455-131_B_F_1865055879 |

**Supplementary Table 3. List of variants covered by SNP chip among 85 suggestive variants**

| rsID | Gene | SNP_chip_name | Probe_ID |
|---|---|---|---|
| rs1405402 | LINC02005-CNTN3 | Human_610_Quad | rs1405402-131_T_R_1857575286 |
| rs1918101 | LINC02005-CNTN3 | Human_610_Quad | rs1918101-131_T_F_1864051902 |
| rs1918101 | LINC02005-CNTN3 | Affymetrix | SNP_A-1869023 |
| rs9855437 | LINC02005-CNTN3 | Affymetrix | SNP_A-4224890 |
| rs941068 | LINC02005-CNTN3 | Affymetrix | SNP_A-2101678 |
| rs6770247 | LINC02005-CNTN3 | Affymetrix | SNP_A-1973279 |
| rs12487132 | LINC02005-CNTN3 | Human_610_Quad | rs12487132-131_B_R_1864233132 |
| rs1958821 | LINC00520-PELI2 | Affymetrix | SNP_A-8422803 |
| rs591012 | TTK | Human_610_Quad | rs591012-131_T_F_1864142653 |
| rs1526709 | LINC02005-CNTN3 | Human_610_Quad | rs1526709-131_B_R_1857613674 |
| rs12639020 | LINC02005-CNTN3 | Affymetrix | SNP_A-2292511 |
| rs239576 | TTK | Affymetrix | SNP_A-8669525 |
| rs152041 | CHP2 | Affymetrix | SNP_A-2169720 |
| rs12447763 | ERN2-CHP2 | Affymetrix | SNP_A-1836328 |
| rs11642116 | CHP2-PRKCB | Human_610_Quad | rs11642116-131_B_R_1864207348 |
| rs2334246 | CHP2-PRKCB | Affymetrix | SNP_A-2245767 |
| rs2152884 | RYR2 | Affymetrix | SNP_A-8604630 |
| rs109592 | CHP2 | Affymetrix | SNP_A-4279623 |
| rs12921822 | RBFOX1 | Affymetrix | SNP_A-1782059 |
| rs9357455 | LINC02530-PHACTR1 | Human_610_Quad | rs9357455-131_B_F_1865055879 |

**Supplementary Table 4. List of variants associated with myocardial infarction in *PHACTR1* gene**

| Study | Variant | Nearest Gene | MAF | P-value | Effect size |
| --- | --- | --- | --- | --- | --- |
| Early-onset AMI associated variant in Korean Genome Project | rs9357455 | LINC02530-PHACTR1 | 0.428 | 9.73E-06 | 0.6658 (OR) |
| Early-onset AMI associated variant in MI consortium | rs12526453 | PHACTR1 | NA | 1.30E-09 | 1.12 (OR) |
| MI associated variant in BBJ | rs9349379 | PHACTR1 | 0.35 | 4.00E-33 | 0.19 (BETA) |
| AMI associated variant in UK biobank | rs9349379 | PHACTR1 | 0.5994 | 1.84E-06 | -0.001266 (BETA) |

**Supplementary Table 5. Functional enrichments in PPI network of the suggestive loci**

| Pathway description | Pathway ID | Pathway Database | false discovery rate | matching proteins in your network (labels) |
|---|---|---|---|---|
| Dissolution of Fibrin Clot | HSA-75205 | REACTOME | 0.0304 | SERPINE1,PLG,PLAUR |
| Complement and coagulation cascades | hsa04610 | KEGG | 0.0093 | SERPINE1,VTN,PLG,PLAUR |
| Cholesterol metabolism | hsa04979 | KEGG | 0.0275 | APOC3,APOA1,LCAT |
| HDL remodeling | HSA-8964058 | REACTOME | 0.0304 | APOC3,APOA1,LCAT |
| RUNX1 regulates genes involved in megakaryocyte differentiation and platelet function | HSA-8936459 | REACTOME | 0.0391 | KMT2C,RBBP5,ASH2L,WDR5 |
| PKMTs methylate histone lysines | HSA-3214841 | REACTOME | 0.0304 | KMT2C,RBBP5,ASH2L,WDR5 |

**Supplementary Table 6. GWAS result belonging to complement and coagulation cascades pathway (hsa04610) in PPI network of the suggestive loci**

| Gene | rsID | P |
|---|---|---|
| PLAUR | rs78631167 | 2.93E-06 |
| VTN | rs181125638 | 0.002132 |
| PLG | rs679814 | 0.003374 |
| SERPINE1 | rs150154382 | 0.04715 |

**Supplementary Table 7. GWAS result belonging to cholesterol metabolism pathway (hsa04979) in PPI network of the suggestive loci**

| Gene | rsID | P |
|---|---|---|
| APOA1 | rs5072 | 8.61E-06 |
| APOC3 | rs5142 | 4.67E-05 |
| LCAT | rs58117942 | 0.006333 |

**Supplementary Table 8. eQTL of the genetic loci for early-onset AMI based on GTEx database**

| rsID | Gene | Gene affected by eQTL | Tissue | Beta | *P* |
|---|---|---|---|---|---|
| rs10232190 | KMT2C | LINC01003 | Heart_Atrial_Appendage | -0.614472 | 1.00E-06 |
| rs10232190 | KMT2C | LINC01003 | Whole_Blood | -0.326114 | 1.16E-06 |
| rs200715251 | GOLGA8T-LINC02249 | CTD-3092A11.3 | Adipose_Visceral_Omentum | -0.486517 | 1.78E-05 |
| rs200715251 | GOLGA8T-LINC02249 | CTD-3092A11.3 | Thyroid | -0.419616 | 6.20E-05 |
| rs200715251 | GOLGA8T-LINC02249 | CTD-3092A11.3 | Whole_Blood | -0.322737 | 8.54E-05 |
| rs200715251 | GOLGA8T-LINC02249 | RP11-540B6.6 | Brain_Cerebellar_Hemisphere | -0.407464 | 9.32E-06 |
| rs200715251 | GOLGA8T-LINC02249 | RP11-540B6.6 | Brain_Cerebellum | -0.405055 | 7.04E-05 |
| rs200715251 | GOLGA8T-LINC02249 | HERC2P10 | Brain_Cerebellum | -0.557554 | 4.43E-05 |
| rs200715251 | GOLGA8T-LINC02249 | RP11-932O9.10 | Skin_Not_Sun_Exposed_Suprapubic | 0.318915 | 0.000113 |
| rs200715251 | GOLGA8T-LINC02249 | RP11-261B23.1 | Testis | -0.533758 | 6.92E-05 |
| rs591012 | TTK | ELOVL4 | Artery_Coronary | -0.2847 | 1.61E-05 |
| rs591012 | TTK | ELOVL4 | Adipose_Visceral_Omentum | -0.231959 | 2.38E-07 |
| rs591012 | TTK | ELOVL4 | Artery_Tibial | -0.29627 | 4.34E-17 |
| rs591012 | TTK | ELOVL4 | Breast_Mammary_Tissue | -0.193601 | 4.63E-06 |
| rs591012 | TTK | ELOVL4 | Adipose_Subcutaneous | -0.263821 | 9.42E-15 |
| rs591012 | TTK | ELOVL4 | Cells_Cultured_fibroblasts | -0.14587 | 1.11E-05 |
| rs591012 | TTK | ELOVL4 | Esophagus_Muscularis | -0.243759 | 7.86E-08 |
| rs591012 | TTK | ELOVL4 | Artery_Aorta | -0.315488 | 4.08E-14 |
| rs591012 | TTK | ELOVL4 | Lung | -0.190903 | 1.96E-06 |
| rs591012 | TTK | ELOVL4 | Muscle_Skeletal | -0.165418 | 0.000137 |
| rs591012 | TTK | ELOVL4 | Nerve_Tibial | -0.33485 | 6.90E-17 |
| rs591012 | TTK | ELOVL4 | Spleen | -0.363117 | 1.26E-06 |

| | | | | | |
|---|---|---|---|---|---|
| rs591012 | TTK | TTK | Adipose_Subcutaneous | -0.16204 | 1.14E-05 |
| rs591012 | TTK | TTK | Cells_Cultured_fibroblasts | -0.066961 | 1.19E-05 |
| rs591012 | TTK | RP11-250B2.5 | Skin_Not_Sun_Exposed_Suprapubic | 0.130621 | 0.000323 |
| rs1614576 | ERN2-CHP2 | PRKCB | Artery_Tibial | 0.305911 | 2.75E-14 |
| rs1614576 | ERN2-CHP2 | PRKCB | Colon_Sigmoid | 0.304845 | 1.31E-08 |
| rs1614576 | ERN2-CHP2 | PRKCB | Adipose_Subcutaneous | -0.125373 | 0.000133 |
| rs1614576 | ERN2-CHP2 | PRKCB | Colon_Transverse | 0.163531 | 2.48E-05 |
| rs1614576 | ERN2-CHP2 | PRKCB | Esophagus_Gastroesophageal_Junction | 0.293643 | 1.18E-08 |
| rs1614576 | ERN2-CHP2 | PRKCB | Esophagus_Muscularis | 0.378902 | 3.33E-18 |
| rs1614576 | ERN2-CHP2 | PRKCB | Artery_Aorta | 0.306525 | 1.22E-09 |
| rs5072 | APOA1-AS | RP11-109L13.1 | Artery_Coronary | -0.976779 | 8.80E-12 |
| rs5072 | APOA1-AS | RP11-109L13.1 | Heart_Left_Ventricle | -0.927041 | 1.67E-13 |
| rs5072 | APOA1-AS | RP11-109L13.1 | Adrenal_Gland | -0.834243 | 1.09E-08 |
| rs5072 | APOA1-AS | RP11-109L13.1 | Adipose_Visceral_Omentum | -0.839566 | 8.57E-14 |
| rs5072 | APOA1-AS | RP11-109L13.1 | Artery_Tibial | -0.965368 | 9.71E-26 |
| rs5072 | APOA1-AS | RP11-109L13.1 | Breast_Mammary_Tissue | -0.992114 | 1.99E-15 |
| rs5072 | APOA1-AS | RP11-109L13.1 | Colon_Sigmoid | -1.02263 | 4.83E-15 |
| rs5072 | APOA1-AS | RP11-109L13.1 | Brain_Cerebellum | -0.919718 | 8.11E-06 |
| rs5072 | APOA1-AS | RP11-109L13.1 | Brain_Cortex | -0.829108 | 8.89E-06 |
| rs5072 | APOA1-AS | RP11-109L13.1 | Brain_Hypothalamus | -1.1296 | 9.62E-08 |
| rs5072 | APOA1-AS | RP11-109L13.1 | Brain_Nucleus_accumbens_basal_ganglia | -0.993106 | 3.40E-07 |

| | | | | | |
|---|---|---|---|---|---|
| rs5072 | APOA1-AS | RP11-109L13.1 | Adipose_Subcutaneous | -1.03097 | 2.60E-25 |
| rs5072 | APOA1-AS | RP11-109L13.1 | Cells_Cultured_fibroblasts | -0.870145 | 1.06E-16 |
| rs5072 | APOA1-AS | RP11-109L13.1 | Cells_EBV-transformed_lymphocytes | -1.02561 | 5.25E-07 |
| rs5072 | APOA1-AS | RP11-109L13.1 | Colon_Transverse | -0.756891 | 5.99E-11 |
| rs5072 | APOA1-AS | RP11-109L13.1 | Esophagus_Gastroesophageal_Junction | -0.856053 | 9.24E-11 |
| rs5072 | APOA1-AS | RP11-109L13.1 | Esophagus_Mucosa | -0.862108 | 1.65E-16 |
| rs5072 | APOA1-AS | RP11-109L13.1 | Esophagus_Muscularis | -0.995614 | 1.15E-21 |
| rs5072 | APOA1-AS | RP11-109L13.1 | Artery_Aorta | -0.856398 | 4.75E-14 |
| rs5072 | APOA1-AS | RP11-109L13.1 | Heart_Atrial_Appendage | -1.04877 | 3.68E-16 |
| rs5072 | APOA1-AS | RP11-109L13.1 | Lung | -0.968593 | 1.56E-20 |
| rs5072 | APOA1-AS | RP11-109L13.1 | Minor_Salivary_Gland | -1.08636 | 7.51E-08 |
| rs5072 | APOA1-AS | RP11-109L13.1 | Muscle_Skeletal | -0.881122 | 3.64E-26 |
| rs5072 | APOA1-AS | RP11-109L13.1 | Nerve_Tibial | -1.00538 | 2.62E-22 |
| rs5072 | APOA1-AS | RP11-109L13.1 | Ovary | -0.759525 | 9.55E-06 |
| rs5072 | APOA1-AS | RP11-109L13.1 | Pituitary | -1.0436 | 1.19E-09 |
| rs5072 | APOA1-AS | RP11-109L13.1 | Prostate | -0.855753 | 3.68E-08 |
| rs5072 | APOA1-AS | RP11-109L13.1 | Skin_Not_Sun_Exposed_Suprapubic | -0.891247 | 1.96E-18 |

| | | | | | |
|---|---|---|---|---|---|
| rs5072 | APOA1-AS | RP11-109L13.1 | Skin_Sun_Exposed_Lower_leg | -1.03835 | 1.12E-29 |
| rs5072 | APOA1-AS | RP11-109L13.1 | Small_Intestine_Terminal_Ileum | -0.884315 | 2.97E-05 |
| rs5072 | APOA1-AS | RP11-109L13.1 | Stomach | -0.757493 | 6.84E-10 |
| rs5072 | APOA1-AS | RP11-109L13.1 | Testis | -1.11959 | 4.98E-15 |
| rs5072 | APOA1-AS | RP11-109L13.1 | Thyroid | -0.861599 | 6.88E-20 |
| rs5072 | APOA1-AS | RP11-109L13.1 | Vagina | -0.854392 | 2.16E-06 |
| rs5072 | APOA1-AS | RP11-109L13.1 | Liver | -0.74007 | 2.19E-06 |
| rs5072 | APOA1-AS | RP11-109L13.1 | Pancreas | -0.955849 | 7.68E-14 |
| rs5072 | APOA1-AS | RP11-109L13.1 | Spleen | -0.877279 | 6.71E-09 |
| rs5072 | APOA1-AS | RP11-109L13.1 | Whole_Blood | -0.858641 | 1.92E-23 |
| rs5072 | APOA1-AS | AP000892.6 | Heart_Left_Ventricle | 0.390127 | 1.93E-07 |
| rs5072 | APOA1-AS | AP000892.6 | Adipose_Visceral_Omentum | 0.329449 | 3.04E-09 |
| rs5072 | APOA1-AS | AP000892.6 | Artery_Tibial | 0.485151 | 4.52E-17 |
| rs5072 | APOA1-AS | AP000892.6 | Colon_Sigmoid | 0.353027 | 5.96E-06 |
| rs5072 | APOA1-AS | AP000892.6 | Adipose_Subcutaneous | 0.418755 | 6.38E-15 |
| rs5072 | APOA1-AS | AP000892.6 | Cells_Cultured_fibroblasts | 0.255656 | 1.61E-05 |
| rs5072 | APOA1-AS | AP000892.6 | Colon_Transverse | 0.251488 | 9.82E-07 |
| rs5072 | APOA1-AS | AP000892.6 | Esophagus_Mucosa | 0.423246 | 5.35E-10 |
| rs5072 | APOA1-AS | AP000892.6 | Esophagus_Muscularis | 0.419377 | 3.47E-08 |
| rs5072 | APOA1-AS | AP000892.6 | Artery_Aorta | 0.560885 | 1.22E-13 |
| rs5072 | APOA1-AS | AP000892.6 | Heart_Atrial_Appendage | 0.50339 | 2.24E-10 |
| rs5072 | APOA1-AS | AP000892.6 | Lung | 0.371691 | 2.73E-10 |
| rs5072 | APOA1-AS | AP000892.6 | Muscle_Skeletal | 0.341494 | 2.71E-15 |
| rs5072 | APOA1-AS | AP000892.6 | Nerve_Tibial | 0.593742 | 4.54E-18 |

| | | | | | |
|---|---|---|---|---|---|
| rs5072 | APOA1-AS | AP000892.6 | Pituitary | 0.484116 | 3.12E-05 |
| rs5072 | APOA1-AS | AP000892.6 | Prostate | 0.34573 | 7.47E-06 |
| rs5072 | APOA1-AS | AP000892.6 | Skin_Not_Sun_Exposed_Suprapubic | 0.434041 | 4.58E-11 |
| rs5072 | APOA1-AS | AP000892.6 | Skin_Sun_Exposed_Lower_leg | 0.480958 | 1.75E-21 |
| rs5072 | APOA1-AS | AP000892.6 | Stomach | 0.325936 | 3.83E-08 |
| rs5072 | APOA1-AS | AP000892.6 | Testis | 0.731808 | 2.32E-12 |
| rs5072 | APOA1-AS | AP000892.6 | Thyroid | 0.39329 | 9.55E-14 |
| rs5072 | APOA1-AS | AP000892.6 | Liver | 0.635195 | 9.45E-09 |
| rs5072 | APOA1-AS | AP000892.6 | Spleen | 0.434425 | 1.81E-06 |
| rs5072 | APOA1-AS | AP000892.6 | Whole_Blood | 0.204766 | 1.09E-09 |
| rs5072 | APOA1-AS | PAFAH1B2 | Cells_Cultured_fibroblasts | 0.119835 | 0.000168 |
| rs5072 | APOA1-AS | PCSK7 | Cells_Cultured_fibroblasts | -0.224947 | 3.96E-11 |
| rs5072 | APOA1-AS | PCSK7 | Lung | -0.157533 | 8.54E-05 |
| rs5072 | APOA1-AS | PCSK7 | Skin_Sun_Exposed_Lower_leg | -0.230749 | 7.50E-07 |
| rs5072 | APOA1-AS | SIDT2 | Lung | 0.301122 | 2.71E-08 |
| rs5072 | APOA1-AS | SIDT2 | Nerve_Tibial | 0.173301 | 0.00012 |
| rs5072 | APOA1-AS | SIDT2 | Thyroid | 0.244384 | 6.78E-07 |
| rs5072 | APOA1-AS | SIDT2 | Whole_Blood | 0.405449 | 1.76E-13 |
| rs5072 | APOA1-AS | AP006216.5 | Testis | 0.457587 | 1.80E-07 |
| rs5072 | APOA1-AS | TAGLN | Testis | 0.3 | 1.72E-05 |
| rs5072 | APOA1-AS | TAGLN | Whole_Blood | 0.55822 | 3.42E-14 |

**Supplementary Table 9. sQTL of the genetic loci for early-onset AMI based on GTEx database**

| rsID | Gene | Gene affected by sQTL | Tissue | Beta | *P* |
|---|---|---|---|---|---|
| rs591012 | TTK | TTK | Cells_Cultured_fibroblasts | 0.416946 | 2.12E-15 |
| rs591012 | TTK | TTK | Cells_EBV-transformed_lymphocytes | 1.00092 | 6.65E-14 |
| rs5072 | APOA1-AS | TAGLN | Adipose_Visceral_Omentum | 0.605286 | 1.39E-08 |
| rs5072 | APOA1-AS | TAGLN | Adrenal_Gland | 0.62809 | 8.28E-06 |
| rs5072 | APOA1-AS | TAGLN | Heart_Left_Ventricle | 0.553836 | 2.43E-06 |
| rs5072 | APOA1-AS | TAGLN | Cells_Cultured_fibroblasts | 0.586946 | 4.54E-12 |
| rs5072 | APOA1-AS | TAGLN | Esophagus_Mucosa | 0.448777 | 6.25E-07 |
| rs5072 | APOA1-AS | TAGLN | Lung | 0.510228 | 1.14E-09 |
| rs5072 | APOA1-AS | TAGLN | Heart_Atrial_Appendage | 0.789882 | 4.34E-12 |
| rs5072 | APOA1-AS | TAGLN | Skin_Not_Sun_Exposed_Suprapubic | -0.336996 | 1.29E-05 |
| rs5072 | APOA1-AS | TAGLN | Skin_Sun_Exposed_Lower_leg | -0.336166 | 9.54E-08 |
| rs5072 | APOA1-AS | PAFAH1B2 | Testis | -0.58628 | 2.01E-06 |
| rs5072 | APOA1-AS | TAGLN | Testis | -0.712932 | 1.14E-07 |
| rs5072 | APOA1-AS | TAGLN | Thyroid | 0.54182 | 4.27E-10 |

**Supplementary Table 10. mQTL of the genetic loci for early-onset AMI based on mQTLdb**

| rsID | Gene | CpG | Gene affected by mQTL | Life stage based on mQTLdb | Beta | *P* | FDR |
|---|---|---|---|---|---|---|---|
| rs2323404 | LINC02005-CNTN3 | cg08015278 | None | adolescence | -0.722257 | 5.99E-28 | 2.07E-21 |
| rs2323404 | LINC02005-CNTN3 | cg08015278 | None | birth | -0.601274 | 4.88E-18 | 1.19E-11 |
| rs2323404 | LINC02005-CNTN3 | cg08015278 | None | childhood | -0.696064 | 9.02E-25 | 2.66E-18 |
| rs2323404 | LINC02005-CNTN3 | cg08015278 | None | middle aged | -0.703125 | 1.03E-24 | 4.38E-18 |
| rs2323404 | LINC02005-CNTN3 | cg08015278 | None | pregnancy | -0.709891 | 1.58E-23 | 6.12E-17 |
| rs10232190 | KMT2C | cg27406678 | NKX6-1 | pregnancy | 0.771899 | 7.41E-08 | 0.043642 |
| rs591012 | TTK | cg24433016 | TTK | adolescence | -0.663543 | 1.07E-62 | 1.14E-55 |
| rs591012 | TTK | cg08355045 | None | adolescence | -0.47803 | 2.38E-30 | 6.99E-24 |
| rs591012 | TTK | cg19323245 | TTK | adolescence | 0.354854 | 5.04E-29 | 1.42E-22 |
| rs591012 | TTK | cg01684562 | TTK | adolescence | -0.314705 | 6.92E-15 | 7.72E-09 |
| rs591012 | TTK | cg21329085 | TTK | adolescence | -0.308465 | 4.81E-14 | 4.98E-08 |
| rs591012 | TTK | cg24433016 | TTK | birth | -0.471394 | 5.22E-30 | 2.18E-23 |
| rs591012 | TTK | cg08355045 | None | birth | -0.473387 | 2.56E-28 | 1.03E-21 |
| rs591012 | TTK | cg19323245 | TTK | birth | 0.246831 | 3.88E-14 | 5.79E-08 |
| rs591012 | TTK | cg01684562 | TTK | birth | -0.279309 | 1.86E-11 | 2.11E-05 |
| rs591012 | TTK | cg21329085 | TTK | birth | -0.336553 | 2.56E-15 | 4.29E-09 |
| rs591012 | TTK | cg24433016 | TTK | childhood | -0.560705 | 6.58E-44 | 4.61E-37 |
| rs591012 | TTK | cg08355045 | None | childhood | -0.526377 | 1.06E-34 | 3.39E-28 |
| rs591012 | TTK | cg19323245 | TTK | childhood | 0.272421 | 4.21E-19 | 6.01E-13 |
| rs591012 | TTK | cg01684562 | TTK | childhood | -0.241488 | 7.07E-10 | 0.000493 |
| rs591012 | TTK | cg21329085 | TTK | childhood | -0.415765 | 1.80E-25 | 4.00E-19 |
| rs591012 | TTK | cg24433016 | TTK | middle aged | -0.606547 | 2.23E-54 | 2.28E-47 |
| rs591012 | TTK | cg08355045 | None | middle aged | -0.476405 | 1.02E-28 | 3.92E-22 |
| rs591012 | TTK | cg19323245 | TTK | middle aged | 0.280078 | 1.47E-17 | 2.42E-11 |
| rs591012 | TTK | cg01684562 | TTK | middle aged | -0.372291 | 3.22E-18 | 5.61E-12 |

| | | | | | | | |
|---|---|---|---|---|---|---|---|
| rs591012 | TTK | cg21329085 | TTK | middle aged | -0.370949 | 4.47E-17 | 7.07E-11 |
| rs591012 | TTK | cg24433016 | TTK | pregnancy | -0.585921 | 2.77E-43 | 1.86E-36 |
| rs591012 | TTK | cg08355045 | None | pregnancy | -0.467062 | 5.01E-25 | 1.48E-18 |
| rs591012 | TTK | cg19323245 | TTK | pregnancy | 0.271854 | 1.96E-14 | 2.36E-08 |
| rs591012 | TTK | cg01684562 | TTK | pregnancy | -0.30438 | 1.94E-13 | 2.15E-07 |
| rs591012 | TTK | cg21329085 | TTK | pregnancy | -0.320726 | 6.86E-14 | 7.85E-08 |
| rs1614576 | ERN2-CHP2 | cg26685404 | PRKCB | adolescence | 0.433301 | 6.66E-24 | 1.25E-17 |
| rs1614576 | ERN2-CHP2 | cg24250393 | PRKCB | adolescence | -0.340804 | 1.97E-14 | 2.27E-08 |
| rs1614576 | ERN2-CHP2 | cg26685404 | PRKCB | birth | 0.487009 | 1.60E-24 | 4.51E-18 |
| rs1614576 | ERN2-CHP2 | cg26685404 | PRKCB | childhood | 0.427774 | 7.91E-22 | 1.34E-15 |
| rs1614576 | ERN2-CHP2 | cg24250393 | PRKCB | childhood | -0.326391 | 1.01E-12 | 9.57E-07 |
| rs1614576 | ERN2-CHP2 | cg26685404 | PRKCB | middle aged | 0.41328 | 1.02E-21 | 2.14E-15 |
| rs1614576 | ERN2-CHP2 | cg24250393 | PRKCB | middle aged | -0.414914 | 2.64E-17 | 4.22E-11 |
| rs1614576 | ERN2-CHP2 | cg26685404 | PRKCB | pregnancy | 0.479934 | 4.67E-23 | 8.97E-17 |
| rs5072 | APOA1-AS | cg11861562 | TAGLN | adolescence | -0.781143 | 9.40E-21 | 5.42E-15 |
| rs5072 | APOA1-AS | cg10130564 | TAGLN | adolescence | -0.721764 | 1.77E-18 | 9.12E-13 |
| rs5072 | APOA1-AS | cg26566898 | TAGLN | adolescence | -0.726552 | 2.32E-18 | 1.18E-12 |
| rs5072 | APOA1-AS | cg15534755 | TAGLN | adolescence | -0.579174 | 3.76E-14 | 1.39E-08 |
| rs5072 | APOA1-AS | cg16524733 | TAGLN | adolescence | -0.604314 | 1.31E-12 | 4.25E-07 |
| rs5072 | APOA1-AS | cg20608306 | SIK3 | adolescence | -0.494754 | 1.50E-12 | 4.82E-07 |
| rs5072 | APOA1-AS | cg05256304 | SIK3 | adolescence | -0.421261 | 1.12E-09 | 0.000271 |
| rs5072 | APOA1-AS | cg11861562 | TAGLN | birth | -0.639393 | 4.57E-14 | 2.33E-08 |
| rs5072 | APOA1-AS | cg10130564 | TAGLN | birth | -0.674137 | 2.49E-15 | 1.40E-09 |
| rs5072 | APOA1-AS | cg26566898 | TAGLN | birth | -0.733155 | 3.53E-17 | 2.31E-11 |
| rs5072 | APOA1-AS | cg15534755 | TAGLN | birth | -0.719382 | 2.38E-18 | 1.65E-12 |
| rs5072 | APOA1-AS | cg16524733 | TAGLN | birth | -0.567948 | 2.37E-11 | 8.88E-06 |

| | | | | | | | |
|---|---|---|---|---|---|---|---|
| rs5072 | APOA1-AS | cg19299755 | None | birth | 0.435344 | 5.56E-08 | 0.013574 |
| rs5072 | APOA1-AS | cg11861562 | TAGLN | childhood | -0.830234 | 8.96E-24 | 5.71E-18 |
| rs5072 | APOA1-AS | cg10130564 | TAGLN | childhood | -0.866268 | 4.49E-27 | 3.38E-21 |
| rs5072 | APOA1-AS | cg26566898 | TAGLN | childhood | -0.818775 | 8.76E-23 | 5.36E-17 |
| rs5072 | APOA1-AS | cg15534755 | TAGLN | childhood | -0.771681 | 4.05E-24 | 2.64E-18 |
| rs5072 | APOA1-AS | cg16524733 | TAGLN | childhood | -0.701469 | 4.12E-17 | 1.82E-11 |
| rs5072 | APOA1-AS | cg20608306 | SIK3 | childhood | -0.450305 | 3.97E-11 | 1.04E-05 |
| rs5072 | APOA1-AS | cg11861562 | TAGLN | middle aged | -0.739044 | 1.21E-15 | 6.68E-10 |
| rs5072 | APOA1-AS | cg10130564 | TAGLN | middle aged | -0.781847 | 1.85E-18 | 1.35E-12 |
| rs5072 | APOA1-AS | cg26566898 | TAGLN | middle aged | -0.776034 | 3.79E-17 | 2.35E-11 |
| rs5072 | APOA1-AS | cg15534755 | TAGLN | middle aged | -0.667174 | 1.91E-15 | 1.04E-09 |
| rs5072 | APOA1-AS | cg16524733 | TAGLN | middle aged | -0.643574 | 2.78E-12 | 1.04E-06 |
| rs5072 | APOA1-AS | cg20608306 | SIK3 | middle aged | -0.396527 | 1.40E-08 | 0.003402 |
| rs5072 | APOA1-AS | cg11861562 | TAGLN | pregnancy | -0.795328 | 4.43E-19 | 3.05E-13 |
| rs5072 | APOA1-AS | cg10130564 | TAGLN | pregnancy | -0.767414 | 2.47E-18 | 1.61E-12 |
| rs5072 | APOA1-AS | cg26566898 | TAGLN | pregnancy | -0.787396 | 6.83E-19 | 4.64E-13 |
| rs5072 | APOA1-AS | cg15534755 | TAGLN | pregnancy | -0.703124 | 1.58E-17 | 9.67E-12 |
| rs5072 | APOA1-AS | cg16524733 | TAGLN | pregnancy | -0.691381 | 2.14E-15 | 1.14E-09 |
| rs5072 | APOA1-AS | cg05256304 | SIK3 | pregnancy | -0.522113 | 8.76E-11 | 2.72E-05 |
| rs12921822 | RBFOX1 | cg06271387 | A2BP1 | middle aged | 0.292434 | 1.64E-09 | 0.003242 |
| rs12921822 | RBFOX1 | cg06271387 | A2BP1 | pregnancy | 0.28483 | 1.54E-09 | 0.002841 |
